# Inverse association between betel quid use and diabetes in rural Bangladesh

**DOI:** 10.1101/2025.07.09.25331184

**Authors:** Kristin K. Sznajder, Mary K. Shenk, Laura Perez, Nurul Alam, Rubhana Raqib, Anjan Kumar, Farjana Haque, Tami Blumenfield, Siobhán M. Cully, Katherine Wander

## Abstract

**Objectives:** Betel quid is used as a mild stimulant in many parts of South and East Asia and the Pacific. In observational studies, its use has been associated with elevated risk for diabetes, but studies in animal models suggest some component(s) of betel quid could reduce risk.

**Methods:** We assessed associations between betel quid use and diabetes (glycated hemoglobin, HbA_1c_ ≥ 6.5%) among a cross-sectional sample of 410 men and 717 non-pregnant women in Matlab, Bangladesh.

**Results:** In multivariable logistic regression, betel quid use was inversely associated with diabetes among men (aOR: 0.45; 95% CI: 0.26, 0.79) but not women (aOR 0.88; 95% CI: 0.51, 1.52). There was a dose-response relationship between frequency of betel quid use and diabetes among men (aOR 0.73; 95% CI: 0.59, 0.89), but not women (aOR 0.96; 95% CI: 0.77, 1.18). Betel quid use was inversely associated with diabetes as an ordinal variable (no diabetes/prediabetes/diabetes) among men (aOR: 0.55; 95% CI: 0.36, 0.82) and women (aOR: 0.66; 95% CI: 0.47, 0.94). Structural equation modeling suggested that some of the inverse association was mediated by food source (market vs. household production).

**Conclusions:** These models support the hypothesis that betel quid use could decrease, rather than increase, risk for diabetes in the Bangladeshi context, particularly among men (who have more frequent betel quid use than women). Heterogeneity in betel quid preparation across settings, multifactorial effects of betel quid use, and a file drawer effect may contribute to differences between these findings and other observational studies.

## Introduction

Betel (or areca) nut is a psychoactive substance (Murphy & Herzog, 2015) used by ∼10% of the world’s population, primarily in South and Southeast Asia and Oceania (Heck et al., 2012; International Agency for Research on Cancer, 2012; C. H. Lee et al., 2011). Preparation of the betel quid varies across contexts, but usually includes the betel nut and calcium hydroxide (slaked lime), often mixed with tobacco, wrapped in a betel leaf (which refers to a climbing vine, distinct from the palm tree from which betel nut is derived), often with spices (e.g., cardamom, clove) or other ingredients included for taste (IARC Working Group on the Evaluation of Carcinogenic Risks to Humans, 2004). This quid is then chewed or held between the teeth and the cheek, allowing mucous membranes of the mouth to absorb the psychoactive ingredient in areca nut, arecoline.

Like many psychoactive substances used in traditional medicine, betel quid seems to have mixed effects on health (summarized in Supporting Information Table S1). Consumers have reported an improved sense of well-being, increased energy, and better digestion (B. J. Boucher & Mannan, 2002). Research with schizophrenia patients found milder symptoms in those who chewed betel quid (Blount, Nguyen, & McDeavitt, 2002; Sullivan, Allen, Otto, Tiobech, & Nero, 2000; Sullivan, Andres D Ch MS, Otto, Miles, & Kydd, 2007). Betel leaf and its component chemicals have been associated with beneficial health effects in humans and animal models, including anticarcinogenic effects, increased antioxidant activity, and antibacterial and antihelminthic properties (Chakraborty & Shah, 2011; Datta, Ghoshdastidar, & Singh, 2011; Padma, Amonkar, & Bhide, 1989; Toprani & Patel, 2013). By contrast, betel quid use has been associated with periodontal disease and damage to teeth, as well as increased risk for cardiovascular disease, hypertension, and oropharyngeal cancer (Anand, Dhingra, Prasad, & Menon, 2014; Chang et al., 2006; W.-Y. Lin et al., 2008; Tseng, 2008). Studies have also found a positive association between betel quid consumption and type 2 diabetes (T2D), and poorer glycemic control (a component of T2D disease etiology) among betel quid users in Taiwan and London (Benjamin, 2001; Mannan, Boucher, & Evans, 2000; Tseng, 2010; Tung et al., 2004).

The link between betel quid consumption and T2D is of particular interest because epidemiologic findings among humans seem to contradict laboratory studies, many of which indicate potential antidiabetic effects of betel quid ingredients in induced animal models (Ahmed et al., 2022; Amudhan & Begum, 2008; Arambewela, Arawwawala, & Ratnasooriya, 2005; P.-L. Huang, Chi, & Liu, 2013; Musdja, Nurdin, & Musir, 2020; Santhakumari, Prakasam, & Pugalendi, 2006). The mechanism(s) by which betel quid use may affect glycemic control and diabetes are summarized in Table 1. Alkaloids from betel quid may inhibit gamma-aminobutyric acid (GABA) receptors, which normally inhibit glucagon secretion (B. J. Boucher & Mannan, 2002), increasing blood glucagon and ultimately blood glucose (W.-Y. Lin et al., 2008). Other studies propose direct toxicity of one or multiple betel quid components to pancreatic beta cells (which secrete insulin) (B. Boucher, Ewen, & Stowers, 1994). An increased risk of obesity associated with betel quid consumption has also been proposed as a potential contributor to the development of T2D in these populations (W. Y. Lin et al., 2009; Mannan et al., 2000). Yet, betel quid consumption has also been associated with appetite suppression (Strickland & Duffield, 1997; Strickland, Veena, Houghton, Stanford, & Kurpad, 2003), which some studies have found to be associated with lower body mass index (BMI) (Suzuki, Jayasena, & Bloom, 2012).

**Table 1.**
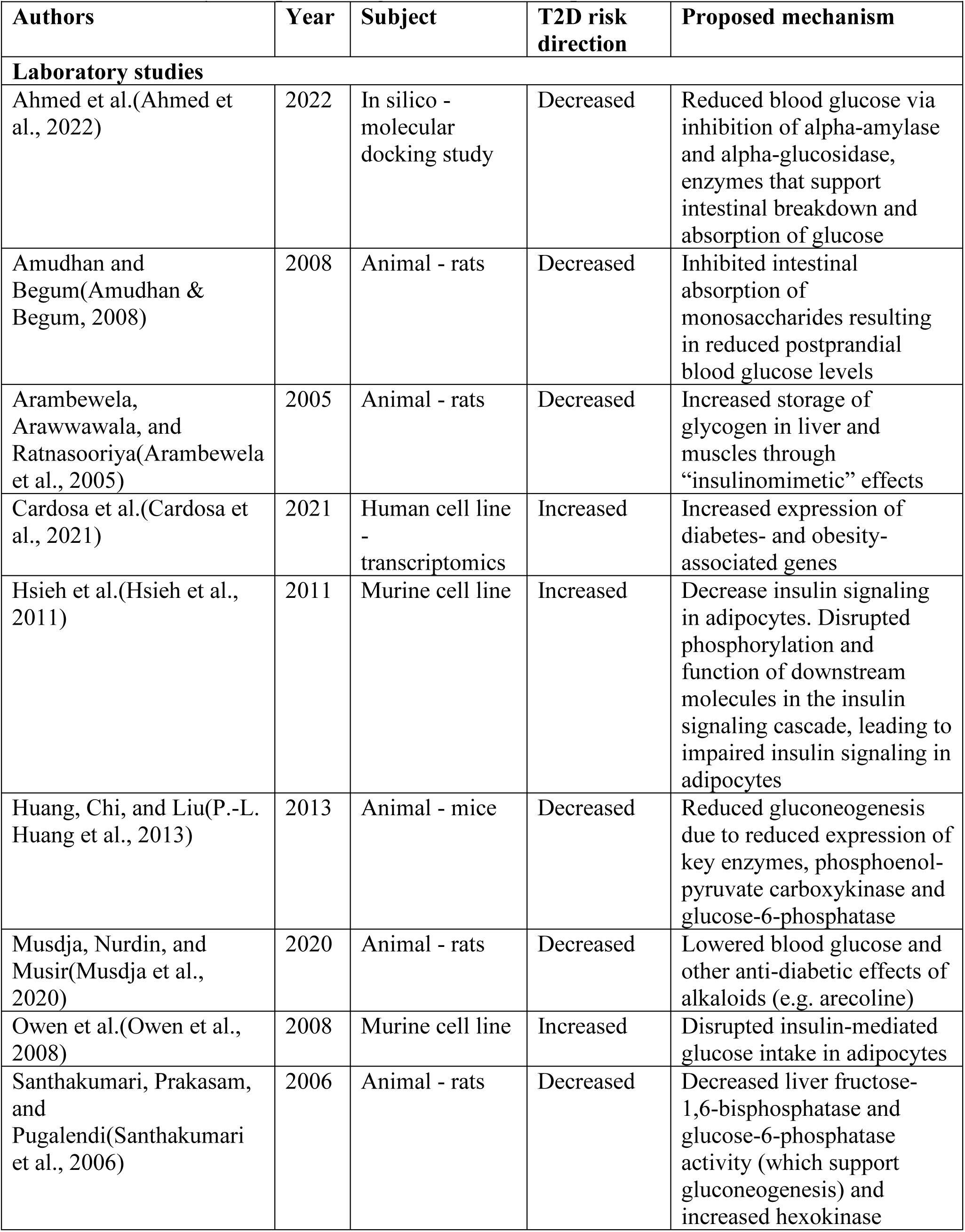
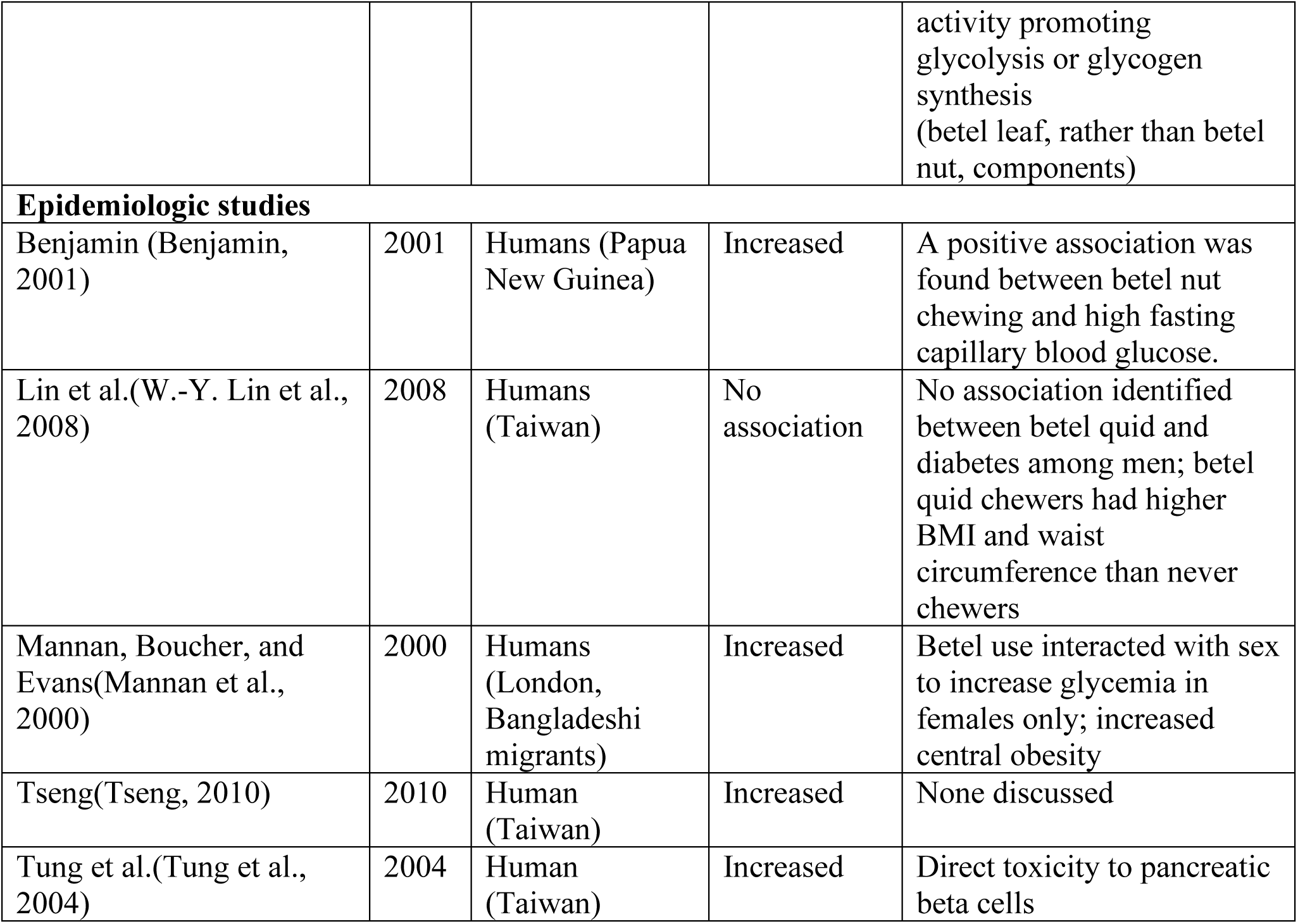
Laboratory and epidemiologic studies of betel quid and diabetes.

Betel quid and T2D
|  |  |  |  |  |
| --- | --- | --- | --- | --- |
| **Authors** | **Year** | **Subject** | **T2D risk direction** | **Proposed mechanism** |
| **Laboratory studies** |  |  |  |  |
| Ahmed et al.(Ahmed et al., 2022) | 2022 | In silico - molecular docking study | Decreased | Reduced blood glucose via inhibition of alpha-amylase and alpha-glucosidase, enzymes that support intestinal breakdown and absorption of glucose |
| Amudhan and Begum(Amudhan & Begum, 2008) | 2008 | Animal - rats | Decreased | Inhibited intestinal absorption of monosaccharides resulting in reduced postprandial blood glucose levels |
| Arambewela, Arawwawala, and Ratnasooriya(Arambewela et al., 2005) | 2005 | Animal - rats | Decreased | Increased storage of glycogen in liver and muscles through “insulinomimetic” effects |
| Cardosa et al.(Cardosa et al., 2021) | 2021 | Human cell line - transcriptomics | Increased | Increased expression of diabetes- and obesity-associated genes |
| Hsieh et al.(Hsieh et al., 2011) | 2011 | Murine cell line | Increased | Decrease insulin signaling in adipocytes. Disrupted phosphorylation and function of downstream molecules in the insulin signaling cascade, leading to impaired insulin signaling in adipocytes |
| Huang, Chi, and Liu(P.-L. Huang et al., 2013) | 2013 | Animal - mice | Decreased | Reduced gluconeogenesis due to reduced expression of key enzymes, phosphoenolpyruvate carboxykinase and glucose-6-phosphatase |
| Musdja, Nurdin, and Musir(Musdja et al., 2020) | 2020 | Animal - rats | Decreased | Lowered blood glucose and other anti-diabetic effects of alkaloids (e.g. arecoline) |
| Owen et al.(Owen et al., 2008) | 2008 | Murine cell line | Increased | Disrupted insulin-mediated glucose intake in adipocytes |
| Santhakumari, Prakasam, and Pugalendi(Santhakumari et al., 2006) | 2006 | Animal - rats | Decreased | Decreased liver fructose-1,6-bisphosphatase and glucose-6-phosphatase activity (which support gluconeogenesis) and increased hexokinase activity promoting glycolysis or glycogen synthesis (betel leaf, rather than betel nut, components) |
| **Epidemiologic studies** |  |  |  |  |
| Benjamin (Benjamin, 2001) | 2001 | Humans (Papua New Guinea) | Increased | A positive association was found between betel nut chewing and high fasting capillary blood glucose. |
| Lin et al.(W.-Y. Lin et al., 2008) | 2008 | Humans (Taiwan) | No association | No association identified between betel quid and diabetes among men; betel quid chewers had higher BMI and waist circumference than never chewers |
| Mannan, Boucher, and Evans(Mannan et al., 2000) | 2000 | Humans (London, Bangladeshi migrants) | Increased | Betel use interacted with sex to increase glycemia in females only; increased central obesity |
| Tseng(Tseng, 2010) | 2010 | Human (Taiwan) | Increased | None discussed |
| Tung et al.(Tung et al., 2004) | 2004 | Human (Taiwan) | Increased | Direct toxicity to pancreatic beta cells |

Betel quid use varies widely across Asia and Oceania, along multiple dimensions, including preparation of the betel nut itself, preparation of the betel quid, and use patterns, so it is not reasonable to expect associations between betel quid use and T2D to be consistent across populations. Indeed, geographic heterogeneity in associations between betel quid use and T2D may help to explain the apparent contrast between observational epidemiologic studies and laboratory-based studies. Thus, we assessed associations between betel quid consumption and diabetes among a cross-sectional sample of men and women living in Matlab, Bangladesh. We further assessed pathways (e.g., BMI) hypothesized to connect betel quid use and T2D.

## Materials and Methods

### Research Setting

We used data collected as part of a larger study on demographic indicators of health and fertility conducted in Matlab, Bangladesh, a densely populated rural area undergoing rapid market integration. Both men and women in this area of Bangladesh commonly chew betel quid, though men chew it more frequently than women, and people from lower socioeconomic status groups use betel quid at higher rates (Wu et al., 2015). Matlab is undergoing a nutrition transition with high and increasing rates of diabetes (S. H. Khan & Talukder, 2013; K. Sznajder et al., 2021).

### Study Design and Participants

A random, age-stratified sample of women was drawn from a population roster of over 230,000 people in the region maintained by the Matlab Health and Demographic Surveillance System (HDSS), which has been operating since the late 1960s by icddr,b (formerly the International Centre for Diarrhoeal Disease Research, Bangladesh) (Alam et al., 2017). Selected women were eligible to participate if they were between 20 and 65 years of age during the first data collection wave in 2010.

Men were eligible to participate if they were married to a participating woman and were present in Matlab at the time of the second data collection wave in 2017; many husbands were not present due to high rates of male labor migration to cities in Bangladesh or abroad (K. Sznajder et al., 2021). This approach ensured a sample that is representative of adult residents of the study area of Matlab, Bangladesh, which for men is disproportionately older and thus potentially in poorer health than a nationally representative sample, due to labor migration. The gender distribution of our sample accurately reflects the actual gender distribution of the population of Matlab (icddr, 2020), as well as many other rural areas of Bangladesh, given the very high rates of male labor outmigration in the county.

All survey, anthropometric, and biomarker data analyzed here were collected in 2017-2018 from 765 women (98% of the 2010 participants who were still alive and continued to reside in the study area), and 499 of their husbands (excluding husbands who were deceased or working as labor migrants outside of Matlab). We excluded participants who reported being pregnant at the time of data collection (N=4), for whom we did not have HbA_1c_ test results (N=114), or for whom we were missing other pertinent variables (N=19) from these analyses.

### Measurements

Data collection instruments were developed and piloted by Shenk and Alam and then implemented by field staff trained at icddr,b. Betel quid use was assessed via survey items that asked participants if they chewed betel quid (yes or no). If yes, participants were asked to report their typical frequency of use, either as times per month or times per day. Based on the reported typical frequency, betel quid use was categorized as none, used monthly, used daily but less than five times per day, and used at least five times per day. Additional variables characterized via surveys included age, years of education, the MacArthur Scale of Subjective Social Status (Adler & Stewart, 2007), occupation (for men only, dichotomized for these analyses as manual laborer or not), social network size (the number of people who the participant interacted with regularly across various social domains including childcare/work assistance, loans of goods or money, and different types of social and emotional support (Shenk et al., 2021)), food security (dichotomized for these analyses as always having enough quality food compared with not always having enough quality food), and food source (all food purchased from the bazaar, where both whole and processed foods are available, compared with some or no food purchased from the bazaar, meaning that food was primarily sourced from the family’s own subsistence activities).

Anthropometry was characterized after survey completion. A stadiometer (Seca 213) was used to measure height in centimeters and a digital scale (Tanita BC 545) was used to measure weight in kilograms (kgs) (Wang & Hui, 2015). BMI was calculated as weight (in kgs) divided by height (in meters) squared and categorized as obese (≥27.5), overweight (23–<27.5), lean (18.5–<23), or underweight (<18.5) according to the World Health Organization’s recommendations for Asian populations (Weir & Jan, 2019). Grip strength was estimated in kgs with a digital hand dynamometer (Takei Grip-D, TKK 5401). Three trials on each hand were included; for these analyses, we used the first trial for the right hand.

A small amount of capillary whole blood was collected from participants by finger stick and immediately evaluated for glycated hemoglobin (HbA_1c_; A1C Now, PTS Diagnostics (Hirst et al., 2017)). Additional drops of whole blood were collected on filter paper (Whatman #903) for dried blood spots (DBS) and allowed to dry for up to 24 hours. DBS were then frozen at −20° C until analysis.

We used 5.7%≤HbA_1c_<6.5% to capture prediabetes and HbA_1c_≥6.5% to capture diabetes and used both binary (no diabetes, diabetes) and ordinal (no diabetes, prediabetes, diabetes) variables to describe diabetes. C-reactive protein (CRP), a biomarker of inflammation, was estimated in DBS at the icddr,b Immunobiology, Nutrition, and Toxicology Laboratory using an enzyme immunoassay kit (BioCheck BC-1119). Kit instructions were modified for DBS as follows: one 3.2 mm disc of DBS specimen was isolated by a manual punch and eluted overnight in assay buffer at 4° C. Eluent was assayed without further dilution. Dilution correction was performed assuming 1.5 µl serum equivalent in a 3.2 mm disc (Brindle, Fujita, Shofer, & O’Connor, 2010). Across 37 plates, the intra-assay coefficient of variation was 5.7% and the inter-assay coefficient of variation was 16.3% (16.7% for a second control specimen included on 27 of these plates). Elevated CRP was defined as >3 mg/l and < 10 mg/l; participants with CRP ≥ 10 mg/l were excluded from models that considered the CRP variable, as concentrations in this range may reflect infectious diseases processes rather than an individual’s characteristic background concentration (Mac Giollabhui et al., 2020; Wander, Brindle, & O’Connor, 2012).

### Statistical Analysis

We estimated logistic regression models of associations between betel quid use and diabetes, and ordinal logistic regression models of associations between betel quid and the ordinal variable of no diabetes, prediabetes, and diabetes. We first estimated crude odds ratios (cOR). We then estimated adjusted odds ratios (aOR) with age, education, MacArthur Scale (for socioeconomic status), and occupation (for men only; most women identified their occupation as housewife) included in models.

Finally, we assessed potential mediators—variables that are conceivably downstream the physiological, behavioral, or social effects of betel quid use, including food security, food source, BMI, CRP, grip strength, and social network size—using structural equation models (SEM). This expansive approach to assessing mediating variables allowed us to assess both direct physiologic effects of betel quid on diabetes risk and indirect effects via food source or quantity, social interactions, and other pathways identifiable with our data.

To evaluate the potential influence of residual confounding, we conducted a sensitivity analysis of our regression models predicting both dichotomous and ordinal diabetes outcomes using E-values (Gaster, Eggertsen, Støvring, Ehrenstein, & Petersen, 2023; VanderWeele & Ding, 2017). The square root of the odds ratios was taken in order to approximate the risk ratio for the E-value calculation. All data were analyzed in SAS 9.4.

### Research Ethics Approval

Procedures were approved by the Pennsylvania State University Institutional Review Board and the Ethical Review Committee at icddr,b, and conformed to the ethical standards of the Declaration of Helsinki and the US Federal Policy for the Protection of Human Subjects. Participants were made aware that study participation was voluntary and provided written consent to participate. HbA_1c_ results (HbA_1c_ in %; diabetic/not), height, weight, BMI, and results of testing not analyzed here were shared with participants (and provided in writing) at the time of data collection.

## Results

Data on diabetes were available for 1127 people, 410 men and 717 non-pregnant women (Table 2). 175 (15.5%) participants were diabetic. Diabetes was more common among men (20.0%) than women (13.0%; p=0.002). Betel quid use (any/none) was similar between men (52.7%) and women (53.4%; p=0.812); however, more men (33.2%) than women (22.2%; p<0.001) reported the highest category of betel quid use (at least five times a day). Participating men were older (56.2 ±12.1 years) than women (49.9 ±12.3 years; p<0.001), had more years of education (4.9 ±4.5 years) than women (4.1 ±3.9 years; p=0.002), had a greater grip strength (26.4 ±10.7 kgs) than women (16.4 ±4.6; p<0.001 kgs), and reported larger social networks (12.7 ±3.9 people) than women (10.4 ±3.8 people; p<0.001). Women reported higher socioeconomic status by MacArthur Ladder (4.6 ±1.8) than men (3.9 ±1.2; p<0.001), were more likely to obtain all their food from the bazaar (63.1%) than men (47.7%; p<0.001), and had higher BMI (23.5 ±4.2 kg/m2) than men (21.5 ±3.6 kg/m2; p<0.001). These gender differences in socioeconomic status likely result from labor migration of younger men to cities in Bangladesh or abroad, including some who would otherwise have participated in our research. Differences in outcomes and key covariates by betel quid use and gender are further described in Supporting Information (Tables S1-S3).

**Table 2.** Sample characteristics N=1127.

| **Table 2. Sample characteristics N=1127** |  |  |  |  |
| --- | --- | --- | --- | --- |
|  | **N (column %) / Mean (SD)** |  |  |  |
|  | Total | Men N=410 | Women N=717 | *p*-value |
| Betel quid use | 599 (53.1) | 216 (52.7) | 383 (53.4) | 0.812 |
| No betel quid use | 528 (46.9) | 194 (47.3) | 334 (46.6) |  |
| Betel quid use $\geq 5$ times per day | 294 (26.2) | 135 (33.2) | 159 (22.2) | <0.001 |
| Betel quid use < 5 times per day | 268 (23.9) | 72 (17.7) | 196 (27.4) |  |
| Betel quid use monthly | 33 (2.9) | 6 (1.5) | 27 (3.8) |  |
| No betel quid use | 528 (47.0) | 194 (47.7) | 334 (46.7) |  |
| Diabetes | 175 (15.5) | 82 (20.0) | 93 (12.9) | <0.001 |
| Prediabetes | 460 (40.8) | 178 (43.4) | 282 (39.3) |  |
| No diabetes | 492 (43.7) | 150 (36.6) | 342 (47.7) |  |
| Age | 52.2 (±12.6) | 56.2 (±12.1) | 49.9 (±12.3) | <0.001 |
| MacArthur Ladder | 4.3 (±1.7) | 3.9 (±1.2) | 4.6 (±1.8) | <0.001 |
| Education | 4.4 (±4.2) | 4.9 (±4.5) | 4.1 (±3.9) | 0.002 |
| No school | 355 (31.8) | 113 (27.8) | 242 (33.9) | 0.028 |
| Up to primary | 385 (34.4) | 137 (33.7) | 248 (34.8) |  |
| More than primary | 378 (33.8) | 156 (38.4) | 222 (31.2) |  |
| Food secure | 752 (67.0) | 280 (68.6) | 472 (66.1) | 0.388 |
| Not food secure | 370 (32.9) | 128 (31.4) | 242 (33.9) |  |
| Food from the bazaar | 646 (57.5) | 194 (47.7) | 452 (63.1) | <0.001 |
| Not all food from bazaar | 477 (42.5) | 213 (52.3) | 264 (36.9) |  |
| Men who are laborers | 218 (54.4) | 218 (54.4) |  |  |
| Men of other occupation | 183 (45.6) | 183 (45.6) |  |  |
| BMI | 22.8 (±4.1) | 21.5 (±3.6) | 23.5 (±4.2) | <0.001 |
| Obese $\geq 27.5$ | 150 (13.4) | 29 (7.1) | 121 (16.9) | <0.001 |
| Overweight 23–<27.5 | 345 (30.7) | 100 (24.5) | 245 (34.3) |  |
| Lean 18.5–<23 | 460 (40.9) | 188 (45.9) | 272 (38.0) |  |
| Underweight <18.5 | 169 (15.0) | 92 (22.5) | 77 (10.8) |  |
| CRP >3–<10 | 240 (22.1) | 78 (19.8) | 162 (23.4) | 0.161 |
| CRP $\leq 3$ | 847 (77.9) | 317 (80.3) | 530 (76.6) | |
| Grip strength (R1) | 19.9 (±7.4) | 26.1 (±7.5) | 16.5 (±4.5) | <0.001 |
| Social network size | 11.2 (±4.0) | 12.7 (±3.9) | 10.4 (±3.8) | <0.001 |
| *Chi-square test for gender difference in means |  |  |  |  |

Betel quid use and gender interacted in their association with diabetes (p<0.001), so all further regression and mediation analyses were stratified by gender. Stratified crude models show an inverse association between betel quid use and diabetes among men (cOR: 0.50; 95% CI: 0.31, 0.82, Table 3, Panel 1) and a positive association among women (cOR: 1.61; 95% CI: 1.02, 2.52, Table 4, Panel 1).

In multivariable models, betel quid use was independently inversely associated with diabetes among men (aOR: 0.45; 95% CI: 0.26, 0.79; Table 3, Panel 2). Further, in ordinal logistic regression using the categorical outcome no diabetes/prediabetes/diabetes, betel quid use was independently inversely associated with diabetes progression among men (aOR: 0.55; 95% CI:0.36, 0.82; Table 3, Panel 2). E-values for associations between betel quid use and dichotomous diabetes (2.24; 95% CI: 1.31, 3.33) and the ordinal diabetes outcome (1.86; 95% CI: 1.19, 2.55) among men suggest that an uncontrolled confounding variable would need to have a moderate to strong association with both betel quid use and diabetes to fully explain the observed associations. Finally, a test for trend to examine a dose-response relationship between the frequency of betel quid use and diabetes supported the finding of an inverse association between diabetes and betel quid use among men (aOR: 0.73; 95% CI: 0.59, 0.89), considering the same variables in Table 3, Panel 2.

**Table 3.**
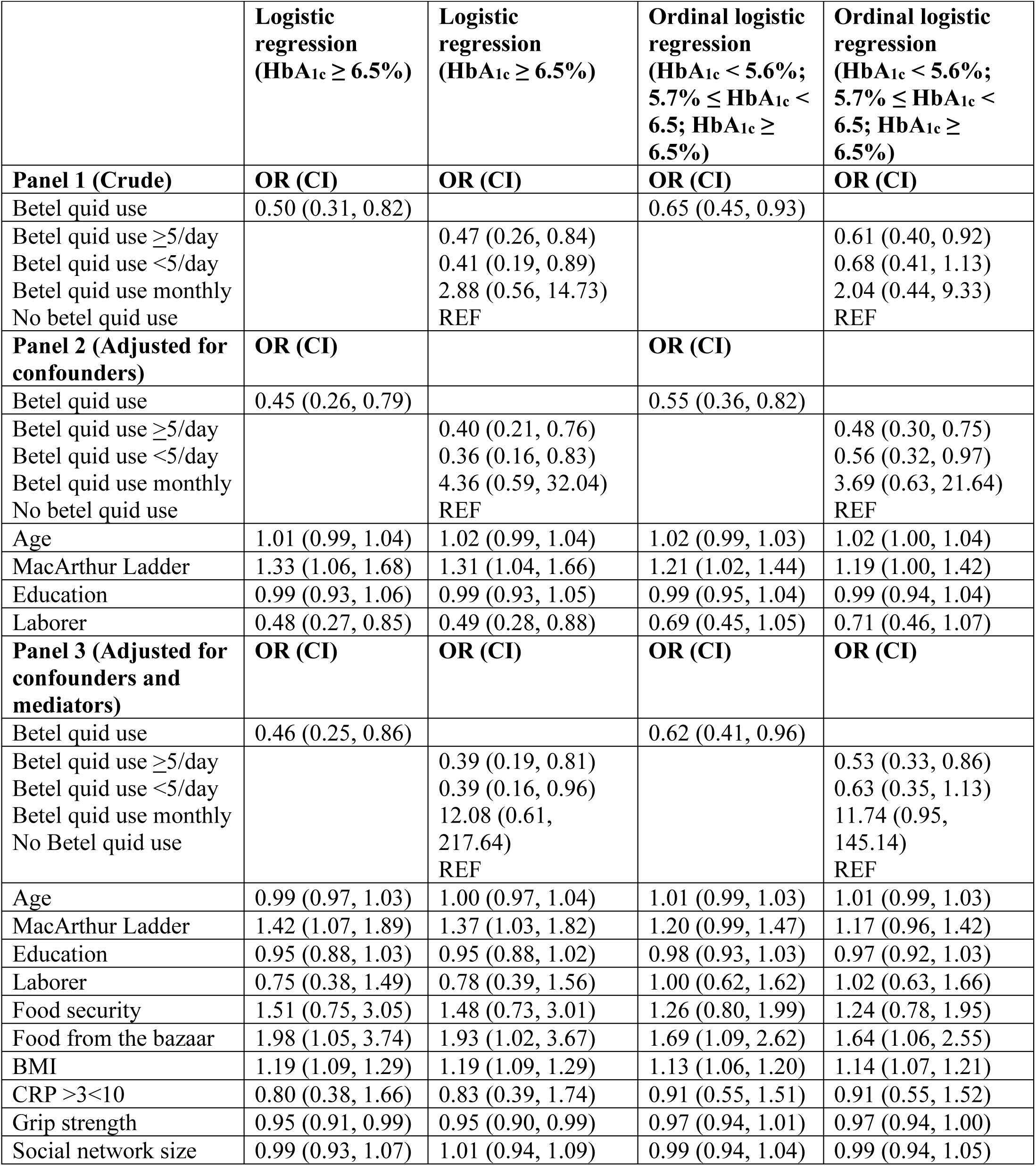
Crude and adjusted logistic regression models of the association between betel quid use and diabetes among men.

|  | Logistic regression<br>(HbA<sub>1c</sub> ≥ 6.5%) | Logistic regression<br>(HbA<sub>1c</sub> ≥ 6.5%) | Ordinal logistic regression<br>(HbA<sub>1c</sub> < 5.6%;<br>5.7% ≤ HbA<sub>1c</sub> < 6.5; HbA<sub>1c</sub> ≥ 6.5%) | Ordinal logistic regression<br>(HbA<sub>1c</sub> < 5.6%;<br>5.7% ≤ HbA<sub>1c</sub> < 6.5; HbA<sub>1c</sub> ≥ 6.5%) |
| --- | --- | --- | --- | --- |
| **Panel 1 (Crude)** | **OR (CI)** | **OR (CI)** | **OR (CI)** | **OR (CI)** |
| Betel quid use | 0.50 (0.31, 0.82) |  | 0.65 (0.45, 0.93) |  |
| Betel quid use ≥5/day |  | 0.47 (0.26, 0.84) |  | 0.61 (0.40, 0.92) |
| Betel quid use <5/day |  | 0.41 (0.19, 0.89) |  | 0.68 (0.41, 1.13) |
| Betel quid use monthly |  | 2.88 (0.56, 14.73) |  | 2.04 (0.44, 9.33) |
| No betel quid use |  | REF |  | REF |
| **Panel 2 (Adjusted for confounders)** | **OR (CI)** |  | **OR (CI)** |  |
| Betel quid use | 0.45 (0.26, 0.79) |  | 0.55 (0.36, 0.82) |  |
| Betel quid use ≥5/day |  | 0.40 (0.21, 0.76) |  | 0.48 (0.30, 0.75) |
| Betel quid use <5/day |  | 0.36 (0.16, 0.83) |  | 0.56 (0.32, 0.97) |
| Betel quid use monthly |  | 4.36 (0.59, 32.04) |  | 3.69 (0.63, 21.64) |
| No betel quid use |  | REF |  | REF |
| Age | 1.01 (0.99, 1.04) | 1.02 (0.99, 1.04) | 1.02 (0.99, 1.03) | 1.02 (1.00, 1.04) |
| MacArthur Ladder | 1.33 (1.06, 1.68) | 1.31 (1.04, 1.66) | 1.21 (1.02, 1.44) | 1.19 (1.00, 1.42) |
| Education | 0.99 (0.93, 1.06) | 0.99 (0.93, 1.05) | 0.99 (0.95, 1.04) | 0.99 (0.94, 1.04) |
| Laborer | 0.48 (0.27, 0.85) | 0.49 (0.28, 0.88) | 0.69 (0.45, 1.05) | 0.71 (0.46, 1.07) |
| **Panel 3 (Adjusted for confounders and mediators)** | **OR (CI)** | **OR (CI)** | **OR (CI)** | **OR (CI)** |
| Betel quid use | 0.46 (0.25, 0.86) |  | 0.62 (0.41, 0.96) |  |
| Betel quid use ≥5/day |  | 0.39 (0.19, 0.81) |  | 0.53 (0.33, 0.86) |
| Betel quid use <5/day |  | 0.39 (0.16, 0.96) |  | 0.63 (0.35, 1.13) |
| Betel quid use monthly |  | 12.08 (0.61, 217.64) |  | 11.74 (0.95, 145.14) |
| No Betel quid use |  | REF |  | REF |
| Age | 0.99 (0.97, 1.03) | 1.00 (0.97, 1.04) | 1.01 (0.99, 1.03) | 1.01 (0.99, 1.03) |
| MacArthur Ladder | 1.42 (1.07, 1.89) | 1.37 (1.03, 1.82) | 1.20 (0.99, 1.47) | 1.17 (0.96, 1.42) |
| Education | 0.95 (0.88, 1.03) | 0.95 (0.88, 1.02) | 0.98 (0.93, 1.03) | 0.97 (0.92, 1.03) |
| Laborer | 0.75 (0.38, 1.49) | 0.78 (0.39, 1.56) | 1.00 (0.62, 1.62) | 1.02 (0.63, 1.66) |
| Food security | 1.51 (0.75, 3.05) | 1.48 (0.73, 3.01) | 1.26 (0.80, 1.99) | 1.24 (0.78, 1.95) |
| Food from the bazaar | 1.98 (1.05, 3.74) | 1.93 (1.02, 3.67) | 1.69 (1.09, 2.62) | 1.64 (1.06, 2.55) |
| BMI | 1.19 (1.09, 1.29) | 1.19 (1.09, 1.29) | 1.13 (1.06, 1.20) | 1.14 (1.07, 1.21) |
| CRP >3<10 | 0.80 (0.38, 1.66) | 0.83 (0.39, 1.74) | 0.91 (0.55, 1.51) | 0.91 (0.55, 1.52) |
| Grip strength | 0.95 (0.91, 0.99) | 0.95 (0.90, 0.99) | 0.97 (0.94, 1.01) | 0.97 (0.94, 1.00) |
| Social network size | 0.99 (0.93, 1.07) | 1.01 (0.94, 1.09) | 0.99 (0.94, 1.04) | 0.99 (0.94, 1.05) |

Among women, betel quid use (any/none) was not associated with diabetes (aOR: 0.88; 95% CI: 0.51, 1.52; Table 4, Panel 2). However, betel quid use was inversely associated with diabetes disease progression among women in ordinal logistic regression (aOR: 0.66; 95% CI: 0.47, 0.94; Table 4, Panel 2). The test for trend (controlling for the same confounding variables) did not reveal an association between diabetes and increasing frequency of betel quid use (aOR: 0.96; 95% CI: 0.77, 1.18).

**Table 4.** Crude and adjusted logistic regression models of the association between betel quid use and diabetes among women.

|  | **Logistic Regression (HbA<sub>1c</sub> ≥ 6.5%)** | **Logistic Regression (HbA<sub>1c</sub> ≥ 6.5)** | **Ordinal Logistic Regression (HbA<sub>1c</sub> < 5.6%; 5.7% ≤ HbA<sub>1c</sub> < 6.5; HbA<sub>1c</sub> ≥ 6.5%)** | **Ordinal Logistic Regression (HbA<sub>1c</sub> < 5.6%; 5.7% ≤ HbA<sub>1c</sub> < 6.5; HbA<sub>1c</sub> ≥ 6.5%)** |
| --- | --- | --- | --- | --- |
| **Panel 1 (Crude)** | **OR (CI)** | **OR (CI)** | **OR (CI)** | **OR (CI)** |
| Betelnut Use | 1.61 (1.02, 2.52) |  | 1.24 (0.94, 1.64) |  |
| Betel quid use ≥5/day |  | 1.49 (0.85, 2.63) |  | 1.19 (0.84, 1.72) |
| Betel quid use <5/day |  | 1.79 (1.07, 2.99) |  | 1.36 (0.98, 1.91) |
| Betel quid use monthly |  | 1.10 (0.32, 3.86) |  | 0.73 (0.34, 1.59) |
| No Betel quid use |  | REF |  | REF |
| **Panel 2 (Adjusted for confounders)** | **OR (CI)** | **OR (CI)** | **OR (CI)** | **OR (CI)** |
| Betelnut use | 0.88 (0.51, 1.52) |  | 0.66 (0.47, 0.94) |  |
| Betel quid use ≥5/day |  | 0.86 (0.45, 1.65) |  | 0.67 (0.44, 1.02) |
| Betel quid use <5/day |  | 0.91 (0.49, 1.68) |  | 0.69 (0.46, 1.04) |
| Betel quid use monthly |  | 0.76 (0.21, 2.75) |  | 0.49 (0.22, 1.09) |
| No Betel quid use |  | REF |  | REF |
| Age | 1.05 (1.02, 1.07) | 1.05 (1.02, 1.07) | 1.04 (1.03, 1.06) | 1.04 (1.03, 1.06) |
| MacArthur Ladder | 1.12 (0.99, 1.27) | 1.12 (0.99, 1.27) | 1.06 (0.98, 1.15) | 1.06 (0.98, 1.15) |
| Education | 0.99 (0.92, 1.07) | 0.99 (0.92, 1.07) | 0.98 (0.94, 1.03) | 0.98 (0.94, 1.03) |
| **Panel 3: (Adjusted for confounders and mediators)** | **OR (CI)** | **OR (CI)** | **OR (CI)** | **OR (CI)** |
| Betelnut Use | 1.03 (0.56, 1.88) |  | 0.74 (0.51, 1.07) |  |
| Betel quid use ≥5/day |  | 0.66 (0.17, 2.55) |  | 0.71 (0.46, 1.12) |
| Betel quid use <5/day |  | 1.17 (0.59, 2.30) |  | 0.85 (0.55, 1.29) |
| Betel quid use monthly |  | 0.97 (0.48, 1.97) |  | 0.44 (0.19, 1.01) |
| No Betel quid use |  | REF |  | REF |
| Age | 1.04 (1.01, 1.07) | 1.04 (1.01, 1.07) | 1.04 (1.02, 1.06) | 1.04 (1.02, 1.06) |
| MacArthur Ladder | 1.06 (0.93, 1.22) | 1.05 (0.91, 1.21) | 1.02 (0.93, 1.12) | 1.01 (0.93, 1.11) |
| Education | 0.94 (0.86, 1.02) | 0.94 (0.86, 1.02) | 0.94 (0.89, 0.99) | 0.94 (0.89, 0.99) |
| Food security | 1.30 (0.73, 2.32) | 1.31 (0.73, 2.34) | 1.07 (0.76, 1.51) | 1.09 (0.77, 1.54) |
| Food from the bazaar | 1.01 (0.59, 1.71) | 0.99 (0.59, 1.69) | 1.21 (0.88, 1.68) | 1.23 (0.89, 1.69) |
| BMI | 1.09 (1.03, 1.17) | 1.09 (1.03, 1.17) | 1.10 (1.06, 1.15) | 1.11 (1.06, 1.15) |
| CRP >3-<10 | 2.23 (1.31, 3.77) | 2.27 (1.34, 3.86) | 1.59 (1.11, 2.30) | 1.62 (1.12, 2.34) |
| Grip strength (R1) | 0.97 (0.92, 1.03) | 0.97 (0.92, 1.03) | 0.99 (0.96, 1.03) | 0.99 (0.96, 1.03) |
| Social Network Size | 1.05 (0.99, 1.12) | 1.06 (0.99, 1.12) | 1.03 (0.99, 1.08) | 1.04 (0.99, 1.08) |

Proportional odds assumptions were met for all ordinal logistic regression models, meaning the logit surfaces were parallel and the OR were constant across outcome categories.

SEM revealed only one potential causal pathway among the mediating variables we considered among men: betel quid use was inversely associated with food from the bazaar (coef: −0.15; p: 0.003) and food from the bazaar was positively associated with diabetes (coef: 0.09; p: 0.083) (Table 5), consistent with the hypothesis that betel quid use decreased men’s intake of bazaar-sourced foods and thus their diabetes risk. The path through food from the bazaar did not fully explain the association between betel quid use and diabetes among men. SEM among women identified only a weak effect suggesting a potential path through food sourced from the bazaar (Table 5).

**Table 5.** Candidate paths between betel quid use and diabetes estimated via structural equation models controlling for pertinent confounders*.

|  |  |  |  |  |  |
| --- | --- | --- | --- | --- | --- |
| Betel -> mediator |  | Mediator | Mediator -> diabetes |  | Mediation effect Y/N |
| Coef | P |  | Coef | P |  |
| **Men** |  |  |  |  |  |
| Direct Path: -0.15 ($P=0.005$) | | | | | |
| -0.02 | 0.656 | BMI<br>Direct Path: (-0.14; $P=0.006$) | 0.19 | <0.001 | N |
| -0.01 | 0.805 | Inflammation (CRP >3-<10mg/l)<br>Direct Path: (-0.15; $P=0.007$) | -0.03 | 0.535 | N |
| 0.06 | 0.209 | Grip Strength<br>Direct Path: (-0.15; $P=0.004$) | -0.09 | 0.142 | N |
| 0.03 | 0.528 | Food security<br>Direct Path: (-0.15; $P=0.004$) | 0.08 | 0.103 | N |
| -0.15 | 0.003 | Food from the bazaar<br>Direct Path: (-0.14; $P=0.012$) | 0.09 | 0.083 | N |
| 0.02 | 0.744 | Social Network Size<br>Direct Path: (-0.15; $P=0.005$) | 0.01 | 0.923 | N |
| **Women** |  |  |  |  |  |
| Direct Path: -0.03 ($P=0.535$) | | | | | |
| -0.05 | 0.244 | BMI<br>Direct Path: (-0.01; $P=0.793$) | 0.17 | <0.001 | N |
| -0.07 | 0.135 | Inflammation (CRP >3-<10mg/l)<br>Direct Path: (-0.00; $P=0.922$) | 0.16 | <0.001 | N |
| 0.02 | 0.578 | Grip Strength<br>Direct Path: (-0.03; $P=0.501$) | -0.02 | 0.624 | N |
| 0.02 | 0.626 | Food security<br>Direct Path: (-0.03; $P=0.503$) | 0.07 | 0.079 | N |
| -0.15 | <0.001 | Food from the bazaar<br>Direct Path: (-0.02; $P=0.587$) | 0.02 | 0.546 | N |
| 0.13 | 0.004 | Social Network Size<br>Direct Path: (-0.03; $P=0.499$) | 0.06 | 0.131 | N |
| *These models controlled for age, MacArthur Ladder, education, and laborer for men and age, MacArthur Ladder, and education for women |  |  |  |  |  |

## Discussion

Our analysis of a large population-based sample from rural Bangladesh suggests that use of betel quid is associated with lower risk for diabetes among men and possibly women. This relationship persisted after a thorough assessment of confounding and mediation. Our analyses suggest these associations are due to direct pathways between betel quid use and diabetes, as well as a potential indirect pathway through lower consumption of bazaar-sourced foods among men. Weak results for women for the same mediator also suggest that more frequent use of betel quid among women might show a similar trend to men.

The gender difference we observed in associations between betel quid use and diabetes has several possible explanations. One is statistical: men are generally at higher risk for T2D compared with women and, in our data, used betel quid more heavily than women; we are thus in a better position to detect the effects of betel use at higher intensity in men (Kautzky-Willer, Leutner, & Harreiter, 2023). The gender difference we observed could also be physiological, if there is dimorphism in an aspect of physiology that is part of the pathway by which betel acts. For example, the gender difference could be due to sex hormones: testosterone (T) has opposite effects on risk for metabolic disorders among men (lower T, higher risk) and women (higher T, higher risk) (Kim & Halter, 2014). Betel quid increases testosterone in mouse models; if it acts similarly on testosterone in humans, this could contribute to the gender difference in associations with diabetes that we report (N.-Y. J. Yang et al., 2004). Finally, the difference could be behavioral or cultural. In Matlab, men often do more physical labor than women, especially in families that grow or raise some or all of their own food. Betel quid might reduce diabetes risk if those who use it have higher physical activity levels (Lo et al., 2016); overall lower levels of physical activity among women in our study population may have prevented them from exhibiting the protective effect we observed among men (Lo et al., 2016). These explanations are not mutually exclusive.

Our findings differ from studies among humans in other settings that support the hypothesis that betel quid use increases risk for T2D (W.-Y. Lin et al., 2008; Liu, Jiesisibieke, Chien, Lee, & Tung, 2024; Mannan et al., 2000; Tseng, 2010; Tung et al., 2004; Yamada, Hara, & Kadowaki, 2013), but accord with some laboratory-based studies of betel quid components in animal models (Amudhan & Begum, 2008; P.-L. Huang et al., 2013; Musdja et al., 2020). Tung and colleagues found a positive association between betel quid use and T2D in a large, population-based study of Taiwanese men with similar controls for confounding as our models, and a dose-response relationship between frequency of use and T2D which supports causal inference (Tung et al., 2004). Mannan and colleagues found a positive association between betel quid use and diabetes among Bangladeshi women, and a slight decrease in glucose concentration among Bangladeshi men, living in East London (Mannan et al., 2000). Among patients with diagnosed diabetes in Bangladesh, Marzan and colleagues found higher random blood glucose among betel quid users than non-users (Al Marzan et al.).

It is difficult to say whether these studies and our findings, taken together, should be seen as contradictory, given the small number of existing studies and the complexity of the potential pathways between betel quid use and T2D. To date, associations between betel quid use and diabetes have been assessed in Taiwan, Papua New Guinea, UK (among Bangladeshi migrants), and now Bangladesh—widely varying social and environmental contexts. It is possible that some component(s) of this context is important to the impact of betel quid use on diabetes risk. Potentially important differences include the way betel quid is typically prepared, and men’s and women’s social roles, diets, frequency of betel quid use, and levels of physical activity. Laboratory studies have identified pathways between betel quid components and metabolic processes that might both increase and decrease diabetes risk (Table 1) (Ahmed et al., 2022; Amudhan & Begum, 2008; Cardosa et al., 2021; Hsieh et al., 2011; P.-L. Huang et al., 2013; Musdja et al., 2020; Owen et al., 2008; Santhakumari et al., 2006). It is thus possible that the net effect of betel quid use increases diabetes risk in one setting and decreases it in another. For this reason, it is crucial for future research to assess how these contents, such as tannins, flavonoids, and triterpenoids, which can have multiple, complex impacts on disease risk (e.g., anti-inflammatory, anti-microbial, and cholesterol-lowering effects), interact with diets, physical activity, and other factors to influence metabolism and diabetes risk (X. Chen, He, & Deng, 2021; Jing, Xiaolan, Yu, Feng, & Haifeng, 2022; Sohag et al., 2022).

Our finding of an inverse association between betel quid use and diabetes in Matlab was unexpected based on the epidemiologic literature. We thus thoroughly interrogated this association via control for a wide range of hypothesized confounders and mediators. Since no confounding or mediating variables fully explained the observed association, it is conceivable that betel quid use protects against disordered metabolism and diabetes in this population.

We may not be the first to document an inverse association between diabetes and betel quid use. A study has been cited in the popular press as documenting a protective effect of betel quid against diabetes in Papua New Guinea (The Syndey Morning Herald Reporter, 2008), though we were unable to locate any peer-reviewed publications or conference proceedings to substantiate this reporting. While it may be appropriate that this study was not published (e.g., if review identified significant flaws), we are nonetheless concerned that a “file drawer effect”—in which null findings or those that are opposite to predictions are less likely to be submitted or accepted for publication—may limit the literature on this topic (Laitin et al., 2021; Rosenthal, 1979). This could inhibit understanding of whether, when, and how betel quid components affect metabolism and T2D risk.

### Limitations

The cross-sectional nature of our study design introduces some limitations. Most notably, we sampled prevalent, rather than incident, cases of diabetes and so may have oversampled cases with long survival times. It is thus possible that the inverse association we observed between betel quid use and diabetes reflects shorter survival of diabetic people who chew betel quid, rather than lower risk for diabetes among people who use betel quid. In addition, reverse causation is possible, such that diabetes causes people to chew less betel quid (rather than betel use reducing diabetes risk). While we cannot dismiss either of these possibilities, the magnitude and consistency of the association we observed among participants suggests neither alone is an adequate explanation.

We found evidence of mediation among women, but were unable to identify a pathway in SEM. This suggests that despite having examined six potential mediators there is some aspect of this effect we have been unable to capture. We anticipate that future research may be able to better unpack this relationship.

In this study, we were unable to distinguish between T2D and Type 1 diabetes (T1D). However, 90-95% of diabetes diagnoses in Bangladesh are T2D, and T1D might often have been fatal in this region until treatment became available in the recent past (Biswas et al., 2019). In addition, no one in the sample reported having T1D or being on insulin. We thus have reason to believe that very few, if any, cases of T1D are included in our sample. Further, our analysis did not consider participants’ report of past diagnosis of diabetes, in favor of HbA_1c_. Results might have differed if past diagnoses were taken into consideration, but this would not have allowed for stratified analysis of the HbA_1c_ result, and reported diagnoses were fairly rare. In addition, we relied on a point of care test to estimate HbA_1c_, rather than a clinical laboratory method, which could have introduced some non-differential error.

Our sample of men may be particularly unhealthy due to the “healthy migrant effect” in which healthier men are more likely to work as labor migrants (Lu, 2008). However, this is unlikely to bias our results in a way that would produce a spurious inverse association between betel quid use and diabetes within our sample of non-migrant men.

Further, it is possible that residual confounding by diet, physical activity, comorbidities, or other unmeasured risk factors could be influencing the associations we observed. Our sensitivity analysis using E-values indicates that unmeasured confounding would need to have a moderate to strong relationship with both the exposure and with a risk ratio equivalent to the E-value, with the lower bound of 1.31 for the dichotomous diabetes outcome and of 1.19 for the ordinal diabetes outcome to fully explain away the observed associations. While it is always possible to have unmeasured confounders in observational research, we considered many important covariates (age, education, MacArthur Scale of Socioeconomic Status, occupation as a proxy for physical activity among men, and potential mediation by food security, food source, BMI, CRP, grip strength, and social network size) and are confident they do not explain our results. We also do not have data on exactly how betel quid was prepared, nor if preparation differed by gender or participants’ duration of use/age at first use. This limits our ability to explain why our findings differ from other publications.

We relied on self-reports and so our findings are vulnerable to bias and error. In particular, participants may have responded to survey items in ways they anticipated would be positively received by the interviewer (e.g., under-reporting betel quid use); biased error or misreporting by diabetes status could have contributed to our observed associations. However, we have no reason to suspect that participants’ recall differed between diabetic and non-diabetic participants.

## Conclusion

We found an inverse association between betel quid use and diabetes among men in Matlab, Bangladesh that is consistent with the hypothesis that betel quid use might protect against diabetes in this context. While our study has some limitations that should be considered when interpreting the results, in light of the contrast between laboratory-based and epidemiologic studies, the inconsistent findings across populations, and the possibility of a file drawer effect in research on this topic, this finding suggests that it is time to more closely examine the effects of betel quid components on metabolism and diabetes risk. A better understanding of the effects of betel quid on metabolic disorders is relevant to several potentially fruitful future research directions including developing diabetes therapeutics, better elucidating diabetes pathobiology, and better understanding widespread use of non-nutritive substances.

## Data Availability

All data produced in the present study are available upon reasonable request to the authors.

## Acknowledgements

The authors would like to thank the study participants. We would also like to acknowledge Taslim Ali and Fayeza Sultana for help with field management, and Sufia Sultana, Tanima Rashid, Shamsun Nahar, Lutfa Begum, Ummehani Akter, Fatema Khatun, Borhan Uddin, and Reyad Hassan for invaluable assistance with data collection. The research was funded through grant number BCS-1839269 from the U.S. National Science Foundation and by the Pennsylvania State University.

## Supplemental Information

**Table S1.**
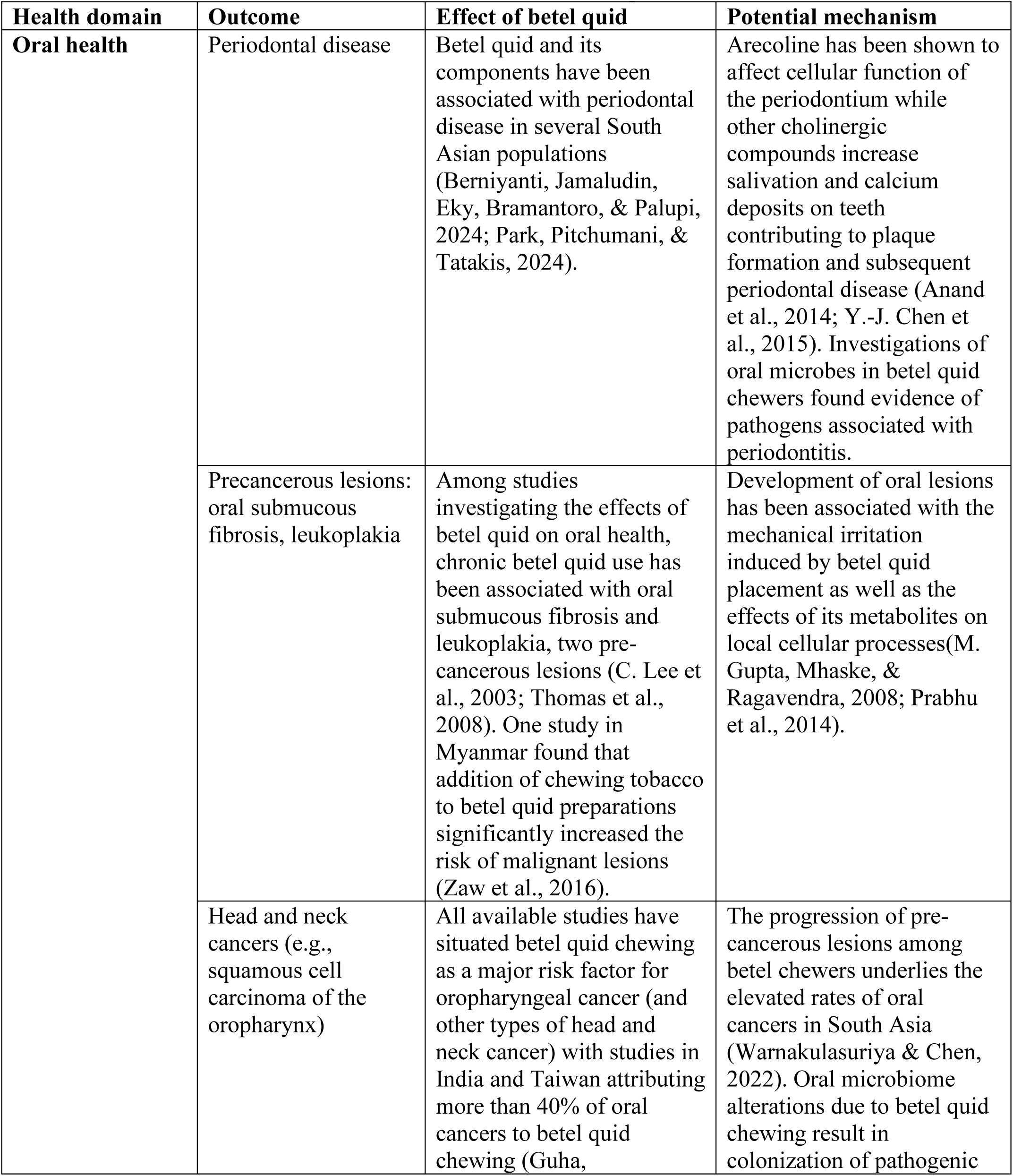
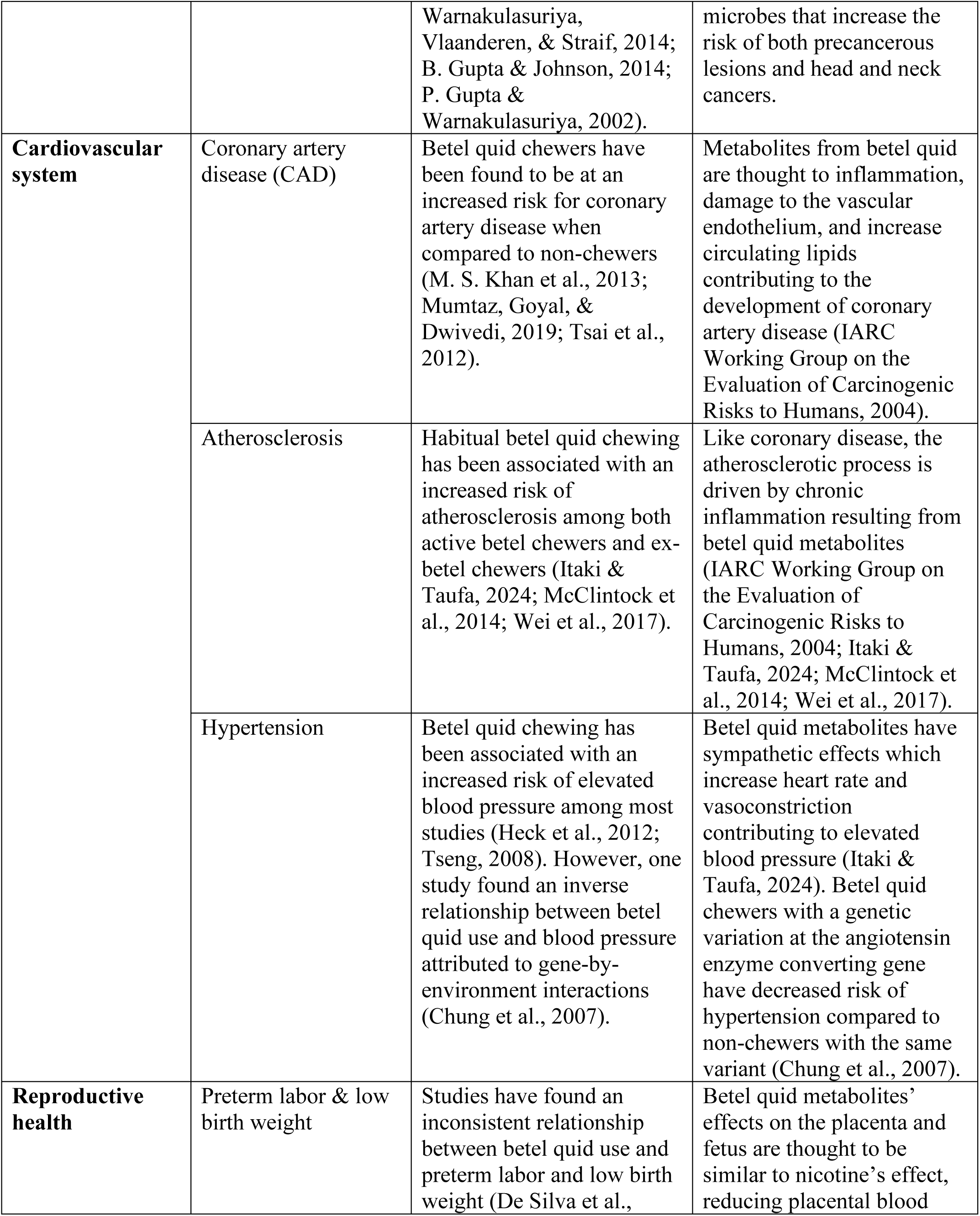
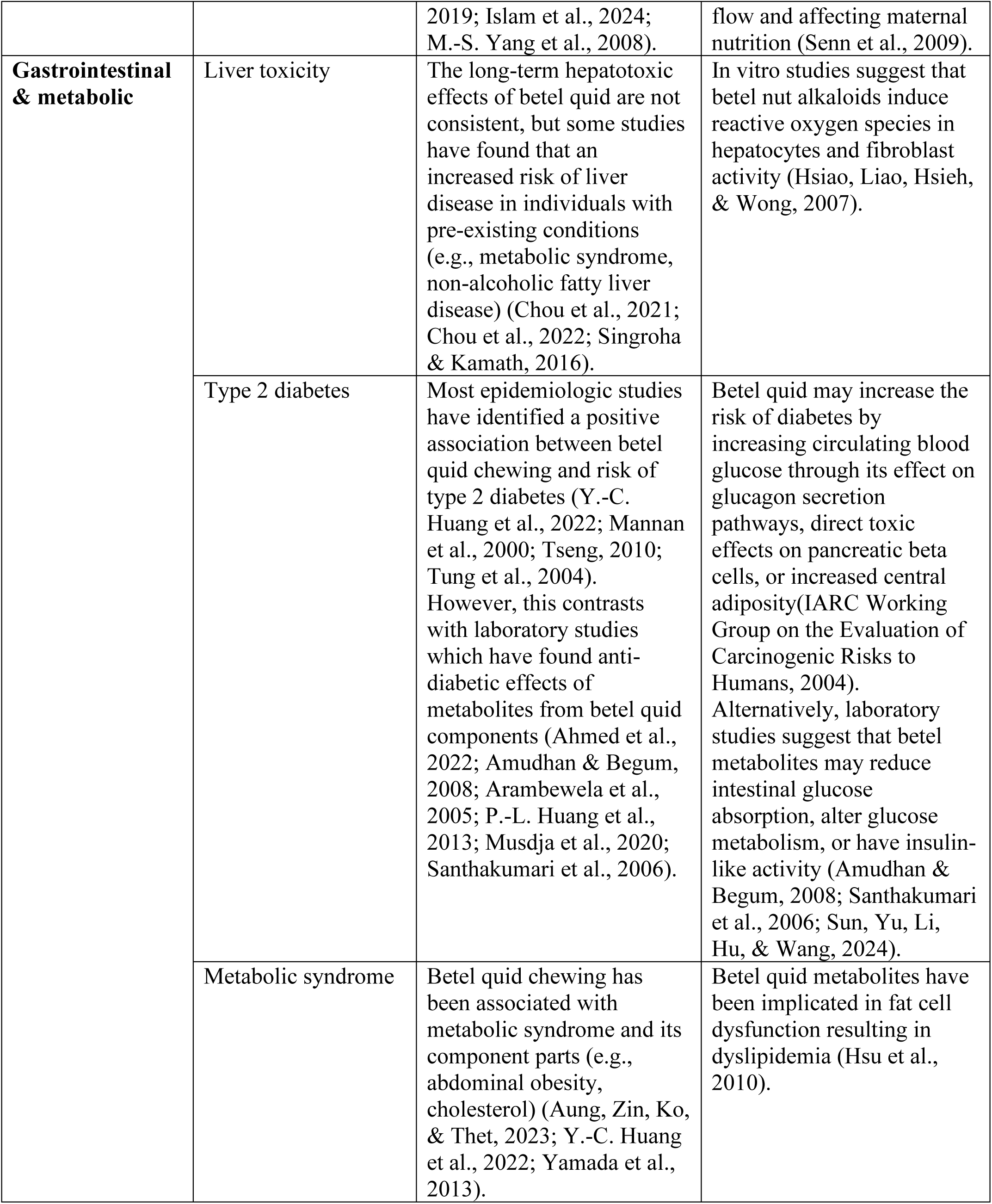
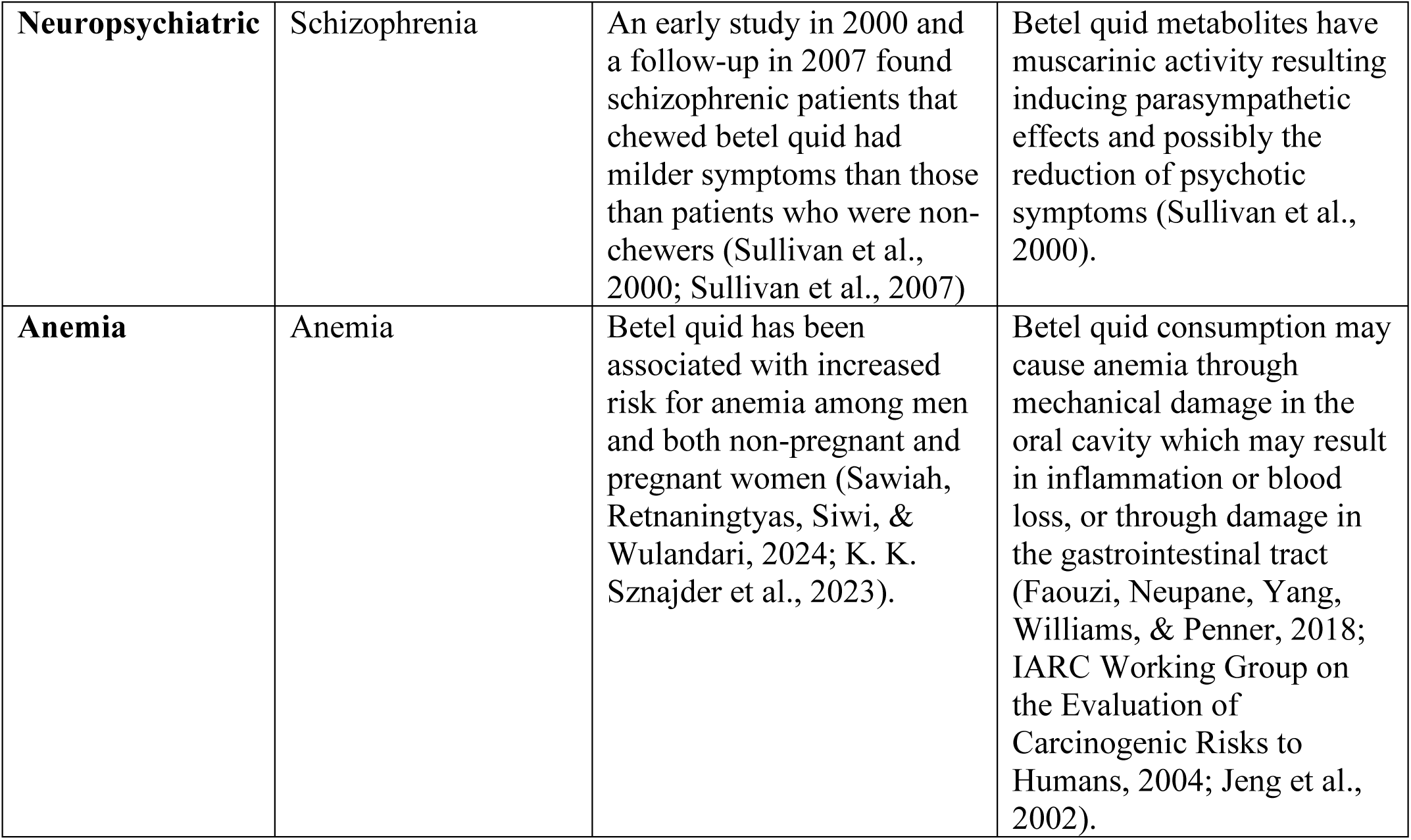
Health outcomes that have been associated with betel quid use.

| **Table S1.** Health outcomes that have been associated with betel quid use. |  |  |  |
| --- | --- | --- | --- |
| **Health domain** | **Outcome** | **Effect of betel quid** | **Potential mechanism** |
| **Oral health** | Periodontal disease | Betel quid and its components have been associated with periodontal disease in several South Asian populations (Berniyanti, Jamaludin, Eky, Bramantoro, & Palupi, 2024; Park, Pitchumani, & Tatakis, 2024). | Arecoline has been shown to affect cellular function of the periodontium while other cholinergic compounds increase salivation and calcium deposits on teeth contributing to plaque formation and subsequent periodontal disease (Anand et al., 2014; Y.-J. Chen et al., 2015). Investigations of oral microbes in betel quid chewers found evidence of pathogens associated with periodontitis. |
|  | Precancerous lesions: oral submucous fibrosis, leukoplakia | Among studies investigating the effects of betel quid on oral health, chronic betel quid use has been associated with oral submucous fibrosis and leukoplakia, two pre-cancerous lesions (C. Lee et al., 2003; Thomas et al., 2008). One study in Myanmar found that addition of chewing tobacco to betel quid preparations significantly increased the risk of malignant lesions (Zaw et al., 2016). | Development of oral lesions has been associated with the mechanical irritation induced by betel quid placement as well as the effects of its metabolites on local cellular processes (M. Gupta, Mhaske, & Ragavendra, 2008; Prabhu et al., 2014). |
|  | Head and neck cancers (e.g., squamous cell carcinoma of the oropharynx) | All available studies have situated betel quid chewing as a major risk factor for oropharyngeal cancer (and other types of head and neck cancer) with studies in India and Taiwan attributing more than 40% of oral cancers to betel quid chewing (Guha, | The progression of pre-cancerous lesions among betel chewers underlies the elevated rates of oral cancers in South Asia (Warnakulasuriya & Chen, 2022). Oral microbiome alterations due to betel quid chewing result in colonization of pathogenic |
|  |  | Warnakulasuriya, Vlaanderen, & Straif, 2014; B. Gupta & Johnson, 2014; P. Gupta & Warnakulasuriya, 2002). | microbes that increase the risk of both precancerous lesions and head and neck cancers. |
| **Cardiovascular system** | Coronary artery disease (CAD) | Betel quid chewers have been found to be at an increased risk for coronary artery disease when compared to non-chewers (M. S. Khan et al., 2013; Mumtaz, Goyal, & Dwivedi, 2019; Tsai et al., 2012). | Metabolites from betel quid are thought to inflammation, damage to the vascular endothelium, and increase circulating lipids contributing to the development of coronary artery disease (IARC Working Group on the Evaluation of Carcinogenic Risks to Humans, 2004). |
|  | Atherosclerosis | Habitual betel quid chewing has been associated with an increased risk of atherosclerosis among both active betel chewers and ex-betel chewers (Itaki & Taufa, 2024; McClintock et al., 2014; Wei et al., 2017). | Like coronary disease, the atherosclerotic process is driven by chronic inflammation resulting from betel quid metabolites (IARC Working Group on the Evaluation of Carcinogenic Risks to Humans, 2004; Itaki & Taufa, 2024; McClintock et al., 2014; Wei et al., 2017). |
|  | Hypertension | Betel quid chewing has been associated with an increased risk of elevated blood pressure among most studies (Heck et al., 2012; Tseng, 2008). However, one study found an inverse relationship between betel quid use and blood pressure attributed to gene-by-environment interactions (Chung et al., 2007). | Betel quid metabolites have sympathetic effects which increase heart rate and vasoconstriction contributing to elevated blood pressure (Itaki & Taufa, 2024). Betel quid chewers with a genetic variation at the angiotensin enzyme converting gene have decreased risk of hypertension compared to non-chewers with the same variant (Chung et al., 2007). |
| **Reproductive health** | Preterm labor & low birth weight | Studies have found an inconsistent relationship between betel quid use and preterm labor and low birth weight (De Silva et al., | Betel quid metabolites' effects on the placenta and fetus are thought to be similar to nicotine's effect, reducing placental blood |
|  |  | 2019; Islam et al., 2024; M.-S. Yang et al., 2008). | flow and affecting maternal nutrition (Senn et al., 2009). |
| **Gastrointestinal & metabolic** | Liver toxicity | The long-term hepatotoxic effects of betel quid are not consistent, but some studies have found that an increased risk of liver disease in individuals with pre-existing conditions (e.g., metabolic syndrome, non-alcoholic fatty liver disease) (Chou et al., 2021; Chou et al., 2022; Singroha & Kamath, 2016). | In vitro studies suggest that betel nut alkaloids induce reactive oxygen species in hepatocytes and fibroblast activity (Hsiao, Liao, Hsieh, & Wong, 2007). |
|  | Type 2 diabetes | Most epidemiologic studies have identified a positive association between betel quid chewing and risk of type 2 diabetes (Y.-C. Huang et al., 2022; Mannan et al., 2000; Tseng, 2010; Tung et al., 2004). However, this contrasts with laboratory studies which have found anti-diabetic effects of metabolites from betel quid components (Ahmed et al., 2022; Amudhan & Begum, 2008; Arambewela et al., 2005; P.-L. Huang et al., 2013; Musdja et al., 2020; Santhakumari et al., 2006). | Betel quid may increase the risk of diabetes by increasing circulating blood glucose through its effect on glucagon secretion pathways, direct toxic effects on pancreatic beta cells, or increased central adiposity(IARC Working Group on the Evaluation of Carcinogenic Risks to Humans, 2004). Alternatively, laboratory studies suggest that betel metabolites may reduce intestinal glucose absorption, alter glucose metabolism, or have insulin-like activity (Amudhan & Begum, 2008; Santhakumari et al., 2006; Sun, Yu, Li, Hu, & Wang, 2024). |
|  | Metabolic syndrome | Betel quid chewing has been associated with metabolic syndrome and its component parts (e.g., abdominal obesity, cholesterol) (Aung, Zin, Ko, & Thet, 2023; Y.-C. Huang et al., 2022; Yamada et al., 2013). | Betel quid metabolites have been implicated in fat cell dysfunction resulting in dyslipidemia (Hsu et al., 2010). |
| **Neuropsychiatric** | Schizophrenia | An early study in 2000 and a follow-up in 2007 found schizophrenic patients that chewed betel quid had milder symptoms than those than patients who were non-chewers (Sullivan et al., 2000; Sullivan et al., 2007) | Betel quid metabolites have muscarinic activity resulting inducing parasympathetic effects and possibly the reduction of psychotic symptoms (Sullivan et al., 2000). |
| **Anemia** | Anemia | Betel quid has been associated with increased risk for anemia among men and both non-pregnant and pregnant women (Sawiah, Retnaningtyas, Siwi, & Wulandari, 2024; K. K. Sznajder et al., 2023). | Betel quid consumption may cause anemia through mechanical damage in the oral cavity which may result in inflammation or blood loss, or through damage in the gastrointestinal tract (Faouzi, Neupane, Yang, Williams, & Penner, 2018; IARC Working Group on the Evaluation of Carcinogenic Risks to Humans, 2004; Jeng et al., 2002). |

**Table S2.** Variables of interest by betel quid use (N=1127)

| **Table S2. Variables of interest by betel quid use (N=1127)** |  |  |  |  |
| --- | --- | --- | --- | --- |
|  | **N (column %)/ Mean (SD)** |  |  |  |
|  | Total | Betel quid<br>use n=599<br>(53.1%) | No Betel quid<br>use n=528<br>(46.9%) | *p*-value |
| Diabetes | 175 (15.5) | 91 (15.2) | 84 (15.9) | 0.944 |
| Prediabetes | 460 (40.8) | 246 (41.1) | 214 (40.5) |  |
| No Diabetes | 492 (43.7) | 262 (43.7) | 230 (43.6) |  |
| Men | 410 (36.4) | 216 (36.1) | 194 (36.7) | 0.812 |
| Women | 717 (63.6) | 383 (63.9) | 334 (63.3) |  |
| Age | 52.2 (±12.6) | 57.4 (±10.2) | 46.3 (±12.4) | <0.001 |
| MacArthur's Ladder Present | 4.3 (±1.7) | 4.3 (±1.7) | 4.3 (±1.7) | 0.864 |
| Education | 4.4 (±4.2) | 2.9 (±3.5) | 6.0 (±4.3) | <0.001 |
| No school | 355 (31.8) | 259 (43.6) | 96 (18.4) | <0.001 |
| Up to Primary | 385 (34.4) | 221 (37.2) | 164 (31.3) |  |
| More than Primary | 378 (33.8) | 114 (19.2) | 264 (50.4) |  |
| Food secure | 752 (67.0) | 395 (66.1) | 357 (68.1) | 0.461 |
| Not food secure | 370 (33.0) | 203 (33.9) | 167 (31.9) |  |
| Food from the bazaar | 646 (57.5) | 309 (51.8) | 337 (64.1) | <0.001 |
| Not all food from bazaar | 477 (42.5) | 288 (48.2) | 189 (35.9) |  |
| Men who are laborers | 218 (54.4) | 128 (60.7) | 90 (47.4) | 0.008 |
| Men of other occupation | 183 (45.6) | 83 (39.3) | 100 (52.6) |  |
| BMI | 22.8 (±4.1) | 22.2 (±4.0) | 23.4 (±4.1) | <0.001 |
| Obese ≥27.5 | 150 (13.4) | 64 (10.7) | 86 (16.4) | <0.001 |
| Overweight 23–<27.5 | 345 (30.7) | 159 (26.6) | 186 (35.4) |  |
| Lean 18.5–<23 | 460 (40.9) | 266 (44.5) | 194 (36.9) |  |
| Underweight<18.5 | 169 (15.0) | 109 (18.2) | 60 (11.4) |  |
| CRP >3-<10 | 240 (22.1) | 114 (19.6) | 126 (24.9) | 0.036 |
| CRP ≤3 | 847 (77.9) | 467 (80.4) | 380 (75.1) | |
| Grip Strength (R1) | 20.1 (±8.8) | 19.0 (±7.3) | 21.2 (±10.2) | <0.001 |
| Social Network Size | 11.2 (±4.0) | 11.2 (±4.0) | 11.3 (±3.9) | 0.754 |

**Table S3.** Bivariate associations between diabetes and variables of interest among men n=410.

|  |  |  |  |  |
| --- | --- | --- | --- | --- |
|  | **N (%) / Mean (SD)** |  |  |  |
|  | Total | Diabetes<br>n=82 (20.0%) | No Diabetes<br>n=328 (80.0%) | *p*-value |
| Betel quid use | 216 (52.7) | 32 (39.0) | 184 (56.1) | 0.006 |
| No Betel quid use | 194 (47.3) | 50 (61.0) | 144 (43.9) |  |
| Betel quid use $\geq 5$ times per day | 135 (33.2) | 19 (23.5) | 116 (35.6) | 0.018 |
| Betel quid use $< 5$ times per day | 78 (19.2) | 12 (14.8) | 66 (20.3) | |
| No Betel quid use | 194 (47.7) | 50 (61.7) | 144 (44.2) |  |
| Age | 56.2 ($\pm 12.1$) | 57.6 ($\pm 11.5$) | 55.9 ($\pm 12.2$) | 0.268 |
| MacArthur's Ladder | 3.9 ($\pm 1.2$) | 4.3 ($\pm 1.2$) | 3.8 ($\pm 1.2$) | $< 0.001$ |
| Education | 4.9 ($\pm 4.5$) | 5.7 ($\pm 5.0$) | 4.7 ($\pm 4.3$) | 0.050 |
| No school | 113 (27.8) | 21 (25.6) | 92 (28.4) | 0.241 |
| Up to Primary | 137 (33.7) | 23 (28.1) | 114 (35.2) |  |
| More than Primary | 156 (38.4) | 38 (46.3) | 118 (36.4) |  |
| Food secure | 280 (68.6) | 67 (81.7) | 213 (65.3) | 0.004 |
| Not food secure | 128 (31.4) | 15 (18.3) | 113 (34.7) |  |
| Food from the bazaar | 194 (47.7) | 50 (61.0) | 144 (44.3) | 0.007 |
| Not all food from bazaar | 213 (52.3) | 32 (39.0) | 181 (55.7) |  |
| Laborer | 218 (54.4) | 28 (34.6) | 190 (59.4) | $< 0.001$ |
| Other occupation | 183 (45.6) | 53 (65.4) | 130 (40.6) |  |
| BMI | 21.5 (3.6) | 23.2 ($\pm 4.4$) | 21.0 ($\pm 3.3$) | $< 0.001$ |
| Obese $\geq 27.5$ | 29 (7.1) | 17 (20.7) | 12 (3.7) | $< 0.001$ |
| Overweight 23–$< 27.5$ | 100 (24.5) | 27 (32.9) | 73 (22.3) | |
| Lean 18.5–$< 23$ | 188 (45.9) | 20 (24.4) | 168 (51.4) | |
| Underweight $< 18.5$ | 92 (22.5) | 18 (21.9) | 74 (22.6) | |
| CRP $> 3$–$< 10$ | 78 (19.8) | 15 (19.5) | 63 (19.8) | 0.948 |
| CRP $\leq 3$ | 317 (80.3) | 62 (80.5) | 255 (80.2) | |
| Grip Strength (R1) | 26.4 ($\pm 10.7$) | 24.7 ($\pm 7.9$) | 26.8 ($\pm 11.3$) | 0.116 |
| Social Network Size | 12.7 ($\pm 3.9$) | 12.8 ($\pm 4.5$) | 12.7 ($\pm 3.9$) | 0.883 |

**Table S4.** Bivariate associations between diabetes and variables of interest among women n=717.

|  |  |  |  |  |
| --- | --- | --- | --- | --- |
|  | **N (%) / Mean (SD)** |  |  |  |
|  | Total | Diabetes n=93 (13.0%) | No Diabetes n=624 (87.0%) | *p*-value |
| Betel quid use | 383 (53.4) | 59 (63.4) | 324 (51.9) | 0.038 |
| No Betel quid use | 334 (46.6) | 34 (36.6) | 300 (48.1) |  |
| Betel quid use $\geq 5$ times per day | 159 (22.2) | 23 (24.7) | 136 (21.8) | 0.100 |
| Betel quid use $< 5$ times per day | 223 (31.2) | 36 (38.7) | 187 (30.0) | |
| No Betel quid use | 334 (46.7) | 34 (36.6) | 300 (48.2) |  |
| Age | 49.9 ($\pm 12.3$) | 55.6 ($\pm 11.2$) | 49.1 ($\pm 12.3$) | $< 0.001$ |
| MacArthur's Ladder Present | 4.6 ($\pm 1.8$) | 5.0 ($\pm 1.8$) | 4.5 ($\pm 1.8$) | 0.011 |
| Education (median in text) | 4.1 ($\pm 3.9$) | 3.3 ($\pm 3.8$) | 4.2 ($\pm 3.9$) | 0.030 |
| No school | 242 (33.9) | 41 (44.6) | 201 (32.4) | 0.018 |
| Up to Primary | 248 (34.8) | 33 (35.9) | 215 (34.7) |  |
| More than Primary | 222 (31.2) | 18 (19.6) | 204 (32.9) |  |
| Food secure | 472 (66.1) | 70 (76.1) | 402 (64.6) | 0.030 |
| Not food secure | 242 (33.9) | 22 (23.9) | 220 (35.4) |  |
| Food from the bazaar | 452 (63.1) | 63 (67.7) | 389 (62.4) | 0.323 |
| Not all food from bazaar | 264 (36.9) | 30 (32.3) | 234 (37.6) |  |
| BMI | 23.5 ($\pm 4.2$) | 25.1 ($\pm 4.1$) | 23.3 ($\pm 4.2$) | $< 0.001$ |
| Obese $\geq 27.5$ | 121 (16.9) | 28 (30.4) | 93 (14.9) | $< 0.001$ |
| Overweight 23–$< 27.5$ | 245 (34.3) | 33 (35.9) | 212 (34.0) | |
| Lean 18.5–$< 23$ | 272 (38.0) | 29 (31.5) | 243 (39.0) | |
| Underweight $< 18.5$ | 77 (10.8) | 2 (2.2) | 75 (12.0) | |
| CRP $> 3$–$< 10$ | 162 (23.4) | 36 (41.9) | 126 (20.8) | $< 0.001$ |
| CRP $\leq 3$ | 530 (76.6) | 50 (58.1) | 480 (79.2) | |
| Grip Strength (R1) | 16.4 ($\pm 4.6$) | 15.7 ($\pm 4.7$) | 16.6 ($\pm 4.5$) | 0.086 |
| Social Network Size | 10.4 ($\pm 3.8$) | 10.9 ($\pm 3.9$) | 10.3 ($\pm 3.7$) | 0.149 |

## References

Adler, N., & Stewart, J. (2007). The MacArthur scale of subjective social status. San Francisco: MacArthur Research Network on SES & Health.

Ahmed, S., Ali, M. C., Ruma, R. A., Mahmud, S., Paul, G. K., Saleh, M. A.,… Rahman, M. M. (2022). Molecular docking and dynamics simulation of natural compounds from betel leaves (Piper betle L.) for investigating the potential inhibition of alpha-amylase and alpha-glucosidase of type 2 diabetes. Molecules, 27(14), 4526.

Al Marzan, A., Hasan, M. S., Islam, M. R., Sakib, M. M., Sifatul, M., Islam, M. S. A.,… Hasan, Z. Impact of betel quid on hyperglycemia among diabetes patients in Bangladesh.

Alam, N., Ali, T., Razzaque, A., Rahman, M., Zahirul Haq, M., Saha, S. K.,… Yunus, M. (2017). Health and demographic surveillance system (HDSS) in Matlab, Bangladesh. International journal of epidemiology, 46(3), 809–816.

Amudhan, M., & Begum, V. (2008). Alpha-glucosidase inhibitory and hypoglycemic activities of Areca catechu extract. Pharmacognosy magazine, 4(15), 223.

Anand, R., Dhingra, C., Prasad, S., & Menon, I. (2014). Betel nut chewing and its deleterious effects on oral cavity. Journal of cancer research and therapeutics, 10(3), 499.

Arambewela, L. S. R., Arawwawala, L., & Ratnasooriya, W. D. (2005). Antidiabetic activities of aqueous and ethanolic extracts of Piper betle leaves in rats. Journal of Ethnopharmacology, 102(2), 239–245.

Aung, A. A., Zin, S. N. S., Ko, A. K., & Thet, A. C. (2023). The Association between Betel Quid Chewing and Metabolic Syndrome Among Urban Adults in Mandalay District of Myanmar. Journal of the ASEAN Federation of Endocrine Societies, 38(2), 50.

Benjamin, A. L. (2001). Community screening for diabetes in the National Capital District, Papua New Guinea: is betelnut chewing a risk factor for diabetes? Papua and New Guinea medical journal, 44(3-4), 101–107. Retrieved from http://europepmc.org/abstract/MED/12422980

Berniyanti, T., Jamaludin, M. B., Eky, Y. E., Bramantoro, T., & Palupi, R. (2024). Duration and frequency of betel quid chewing affects periodontitis severity and life quality of people in Tanini Village, Kupang, Indonesia. International Journal of Dental Hygiene, 22(1), 229–235.

Biswas, T., Townsend, N., Islam, M. S., Islam, M. R., Gupta, R. D., Das, S. K., & Al Mamun, A. (2019). Association between socioeconomic status and prevalence of non-communicable diseases risk factors and comorbidities in Bangladesh: findings from a nationwide cross-sectional survey. BMJ open, 9(3), e025538.

Blount, P. J., Nguyen, C. D., & McDeavitt, J. T. (2002). Clinical use of cholinomimetic agents: a review. The Journal of head trauma rehabilitation, 17(4), 314–321.

Boucher, B., Ewen, S., & Stowers, J. (1994). Betel nut (Areca catechu) consumption and the induction of glucose intolerance in adult CD1 mice and in their F1 and F2 offspring. Diabetologia, 37(1), 49–55.

Boucher, B. J., & Mannan, N. (2002). Metabolic effects of the consumption of Areca catechu. Addiction biology, 7(1), 103–110.

Brindle, E., Fujita, M., Shofer, J., & O’Connor, K. A. (2010). Serum, plasma, and dried blood spot high-sensitivity C-reactive protein enzyme immunoassay for population research. Journal of immunological methods, 362(1-2), 112–120.

Cardosa, S. R., Ogunkolade, B. W., Lowe, R., Savage, E., Mein, C. A., Boucher, B. J., & Hitman, G. A. (2021). Areca catechu-(Betel-nut)-induced whole transcriptome changes in a human monocyte cell line that may have relevance to diabetes and obesity; a pilot study. BMC endocrine disorders, 21(1), 1–11.

Chakraborty, D., & Shah, B. (2011). Antimicrobial, antioxidative and antihemolytic activity of Piper betel leaf extracts. International Journal of Pharmacy and Pharmaceutical Sciences, 3(3), 192–199.

Chang, W., Hsiao, C., Chang, H., Lan, T., Hsiung, C., Shih, Y., & Tai, T. (2006). Betel nut chewing and other risk factors associated with obesity among Taiwanese male adults. International journal of obesity, 30(2), 359–363.

Chen, X., He, Y., & Deng, Y. (2021). Chemical composition, pharmacological, and toxicological effects of betel nut. Evidence-Based Complementary and Alternative Medicine, 2021.

Chen, Y.-J., Lee, S.-S., Huang, F.-M., Yu, H.-C., Tsai, C.-C., & Chang, Y.-C. (2015). Effects of arecoline on cell growth, migration, and differentiation in cementoblasts. Journal of Dental Sciences, 10(4), 388–393.

Chou, Y.-T., Li, C.-H., Sun, Z.-J., Shen, W.-C., Yang, Y.-C., Lu, F.-H.,… Wu, J.-S. (2021). A positive relationship between betel nut chewing and significant liver fibrosis in NAFLD subjects, but not in non-NAFLD ones. Nutrients, 13(3), 914.

Chou, Y.-T., Sun, Z.-J., Shen, W.-C., Yang, Y.-C., Lu, F.-H., Chang, C.-J.,… Wu, J.-S. (2022). Cumulative betel quid chewing and the risk of significant liver fibrosis in subjects with and without metabolic syndrome. Frontiers in Nutrition, 9, 765206.

Chung, F.-M., Shieh, T.-Y., Yang, Y.-H., Chang, D.-M., Shin, S.-J., Tsai, J. C.-R.,… Lee, Y.-J. (2007). The role of angiotensin-converting enzyme gene insertion/deletion polymorphism for blood pressure regulation in areca nut chewers. Translational research, 150(1), 58–65.

Datta, A., Ghoshdastidar, S., & Singh, M. (2011). Antimicrobial property of Piper betel leaf against clinical isolates of bacteria. International Journal of Pharma Sciences and Research, 2(3), 104–109.

De Silva, M., Panisi, L., Brownfoot, F. C., Lindquist, A., Walker, S. P., Tong, S., & Hastie, R. (2019). Systematic review of areca (betel nut) use and adverse pregnancy outcomes. International Journal of Gynecology & Obstetrics, 147(3), 292–300.

Faouzi, M., Neupane, R. P., Yang, J., Williams, P., & Penner, R. (2018). Areca nut extracts mobilize calcium and release pro-inflammatory cytokines from various immune cells. Scientific reports, 8(1), 1075.

Gaster, T., Eggertsen, C. M., Støvring, H., Ehrenstein, V., & Petersen, I. (2023). Quantifying the impact of unmeasured confounding in observational studies with the E value. BMJ medicine, 2(1), e000366.

Guha, N., Warnakulasuriya, S., Vlaanderen, J., & Straif, K. (2014). Betel quid chewing and the risk of oral and oropharyngeal cancers: a meta-analysis with implications for cancer control. International Journal of Cancer, 135(6), 1433–1443.

Gupta, B., & Johnson, N. W. (2014). Systematic review and meta-analysis of association of smokeless tobacco and of betel quid without tobacco with incidence of oral cancer in South Asia and the Pacific. PloS one, 9(11), e113385.

Gupta, M., Mhaske, S., & Ragavendra, R. (2008). Oral submucous fibrosis: current concepts in etiopathogenesis.

Gupta, P., & Warnakulasuriya, S. (2002). Global epidemiology of areca nut usage. Addiction biology, 7(1), 77–83.

Heck, J. E., Marcotte, E. L., Argos, M., Parvez, F., Ahmed, A., Islam, T.,… Chen, Y. (2012). Betel quid chewing in rural Bangladesh: prevalence, predictors and relationship to blood pressure. International journal of epidemiology, 41(2), 462–471.

Hirst, J. A., McLellan, J. H., Price, C. P., English, E., Feakins, B. G., Stevens, R. J., & Farmer, A. J. (2017). Performance of point-of-care HbA1c test devices: implications for use in clinical practice – a systematic review and meta-analysis. Clinical Chemistry and Laboratory Medicine (CCLM), 55(2), 167–180. doi:doi:10.1515/cclm-2016-0303

Hsiao, T.-J., Liao, H.-W. C., Hsieh, P.-S., & Wong, R.-H. (2007). Risk of betel quid chewing on the development of liver cirrhosis: a community-based case-control study. Annals of Epidemiology, 17(6), 479–485.

Hsieh, T.-J., Hsieh, P.-C., Wu, M.-T., Chang, W.-C., Hsiao, P.-J., Lin, K.-D.,… Shin, S.-J. (2011). Betel nut extract and arecoline block insulin signaling and lipid storage in 3T3-L1 adipocytes. Cell Biology and Toxicology, 27, 397–411.

Hsu, H.-F., Tsou, T.-C., Chao, H.-R., Shy, C.-G., Kuo, Y.-T., Tsai, F.-Y.,… Ko, Y.-C. (2010). Effects of arecoline on adipogenesis, lipolysis, and glucose uptake of adipocytes—A possible role of betel-quid chewing in metabolic syndrome. Toxicology and applied pharmacology, 245(3), 370–377.

Huang, P.-L., Chi, C.-W., & Liu, T.-Y. (2013). Areca nut procyanidins ameliorate streptozocin-induced hyperglycemia by regulating gluconeogenesis. Food and chemical toxicology, 55, 137–143.

Huang, Y.-C., Geng, J.-H., Wu, P.-Y., Huang, J.-C., Chen, S.-C., Chang, J.-M., & Chen, H.-C. (2022). Betel nut chewing increases the risk of metabolic syndrome and its components in a large Taiwanese population follow-up study category: original investigation. Nutrients, 14(5), 1018.

IARC Working Group on the Evaluation of Carcinogenic Risks to Humans. (2004). Betel-quid and areca-nut chewing and some areca-nut derived nitrosamines. IARC Monogr Eval Carcinog Risks Hum, 85(1).

icddr, b. (2020). Health and Demographic Surveillance System–Matlab v.53. Retrieved from Dhaka: icddr,b.:

International Agency for Research on Cancer. (2012). Betel quid and area nut. In IARC Working Group on the Evaluation of Carcinogenic Risks to Humans. Personal Habits and Indoor Combustions (Vol. IARC Monographs on the Evaluation of Carcinogenic Risks to Humans, No. 100E). Lyon, France: Available from: https://www.ncbi.nlm.nih.gov/books/NBK304393/.

Islam, M. R., Aktar, S., Pervin, J., Rahman, S. M., Rahman, M., Rahman, A., & Ekström, E.-C. (2024). Maternal betel quid use during pregnancy and child growth: a cohort study from rural Bangladesh. Global health action, 17(1), 2375829.

Itaki, R., & Taufa, S. (2024). Association between habitual betel quid chewing and risk of adverse cardiovascular outcomes: A systematic review. Tropical Medicine & International Health.

Jeng, J.-H., Chen, S.-Y., Liao, C.-H., Tung, Y.-Y., Lin, B.-R., Hahn, L.-J., & Chang, M.-C. (2002). Modulation of platelet aggregation by areca nut and betel leaf ingredients: roles of reactive oxygen species and cyclooxygenase. Free Radical Biology and Medicine, 32(9), 860–871.

Jing, W., Xiaolan, C., Yu, C., Feng, Q., & Haifeng, Y. (2022). Pharmacological effects and mechanisms of tannic acid. Biomedicine & Pharmacotherapy, 154, 113561.

Kautzky-Willer, A., Leutner, M., & Harreiter, J. (2023). Sex differences in type 2 diabetes. Diabetologia, 66(6), 986–1002. doi:10.1007/s00125-023-05891-x

Khan, M. S., Bawany, F. I., Ahmed, M. U., Hussain, M., Khan, A., & Lashari, M. N. (2013). Betel nut usage is a major risk factor for coronary artery disease. Global journal of health science, 6(2), 189.

Khan, S. H., & Talukder, S. H. (2013). Nutrition transition in Bangladesh: is the country ready for this double burden. obesity reviews, 14, 126–133.

Kim, C., & Halter, J. B. (2014). Endogenous sex hormones, metabolic syndrome, and diabetes in men and women. Current cardiology reports, 16, 1–12.

Laitin, D. D., Miguel, E., Alrababa’h, A., Bogdanoski, A., Grant, S., Hoeberling, K.,… Weinstein, J. (2021). Reporting all results efficiently: A RARE proposal to open up the file drawer. Proceedings of the National Academy of Sciences, 118(52), e2106178118.

Lee, C., Ko, Y., Huang, H., Chao, Y., Tsai, C., Shieh, T., & Lin, L. (2003). The precancer risk of betel quid chewing, tobacco use and alcohol consumption in oral leukoplakia and oral submucous fibrosis in southern Taiwan. British journal of cancer, 88(3), 366–372.

Lee, C. H., Ko, A. M. S., Warnakulasuriya, S., Yin, B. L., Zain, R. B., Ibrahim, S. O.,… Utomo, B. (2011). Intercountry prevalences and practices of betel-quid use in south, southeast and eastern Asia regions and associated oral preneoplastic disorders: an international collaborative study by Asian betel-quid consortium of south and east Asia. International Journal of Cancer, 129(7), 1741–1751.

Lin, W.-Y., Chiu, T.-Y., Lee, L.-T., Lin, C.-C., Huang, C.-Y., & Huang, K.-C. (2008). Betel nut chewing is associated with increased risk of cardiovascular disease and all-cause mortality in Taiwanese men. The American journal of clinical nutrition, 87(5), 1204–1211.

Lin, W. Y., Pi-Sunyer, F. X., Liu, C. S., Li, T. C., Li, C. I., Huang, C. Y., & Lin, C. C. (2009). Betel nut chewing is strongly associated with general and central obesity in Chinese male middle-aged adults. Obesity, 17(6), 1247–1254.

Liu, W.-Y., Jiesisibieke, Z. L., Chien, C.-W., Lee, E. K.-L., & Tung, T.-H. (2024). Diabetes mellitus associated with areca nut usage: A systematic review and meta-analysis. Preventive medicine, 107922.

Lo, F. E., Lu, P. J., Tsai, M. K., Lee, J. H., Wen, C., Wen, C. P.,… Lyu, S. Y. (2016). The role of physical activity in harm reduction among betel quid chewers from a prospective cohort of 419,378 individuals. PloS one, 11(4), e0152246.

Lu, Y. (2008). Test of the ‘healthy migrant hypothesis’: a longitudinal analysis of health selectivity of internal migration in Indonesia. Social science & medicine, 67(8), 1331–1339.

Mac Giollabhui, N., Ellman, L. M., Coe, C. L., Byrne, M. L., Abramson, L. Y., & Alloy, L. B. (2020). To exclude or not to exclude: Considerations and recommendations for C-reactive protein values higher than 10 mg/L. Brain, behavior, and immunity, 87, 898.

Mannan, N., Boucher, B., & Evans, S. (2000). Increased waist size and weight in relation to consumption of Areca catechu (betel-nut); a risk factor for increased glycaemia in Asians in east London. British Journal of Nutrition, 83(3), 267–275.

McClintock, T. R., Parvez, F., Wu, F., Wang, W., Islam, T., Ahmed, A.,… Desvarieux, M. (2014). Association between betel quid chewing and carotid intima-media thickness in rural Bangladesh. International journal of epidemiology, 43(4), 1174–1182.

Mumtaz, S. M., Goyal, R. K., & Dwivedi, S. (2019). Adverse cardiovascular effects of betel nut. MGM Journal of Medical Sciences, 6(4), 171–174.

Murphy, K. L., & Herzog, T. A. (2015). Sociocultural factors that affect chewing behaviors among betel nut chewers and ex-chewers on Guam. Hawai’i Journal of Medicine & Public Health, 74(12), 406.

Musdja, M. Y., Nurdin, A., & Musir, A. (2020). Antidiabetic effect and glucose tolerance of areca nut (Areca catechu) seed ethanol extract on alloxan-induced diabetic male rats. Paper presented at the IOP Conference Series: Earth and Environmental Science.

Owen, P. L., Martineau, L. C., Caves, D., Haddad, P. S., Matainaho, T., & Johns, T. (2008). Consumption of guava (Psidium guajava L) and noni (Morinda citrifolia L) may protect betel quid-chewing Papua New Guineans against diabetes. Asia Pacific Journal of Clinical Nutrition, 17(4).

Padma, P., Amonkar, A., & Bhide, S. (1989). Antimutagenic effects of betel leaf extract against the mutagenicity of two tobacco-specific N-nitrosamines. Mutagenesis, 4(2), 154–156.

Park, J. V., Pitchumani, P. K., & Tatakis, D. N. (2024). Periodontitis presenting among betel quid users: A case series. Clinical Advances in Periodontics.

Prabhu, R. V., Prabhu, V., Chatra, L., Shenai, P., Suvarna, N., & Dandekeri, S. (2014). Areca nut and its role in oral submucous fibrosis. Journal of clinical and experimental dentistry, 6(5), e569.

Rosenthal, R. (1979). The file drawer problem and tolerance for null results. Psychological bulletin, 86(3), 638.

Santhakumari, P., Prakasam, A., & Pugalendi, K. V. (2006). Antihyperglycemic activity of Piper betle leaf on streptozotocin-induced diabetic rats. Journal of medicinal food, 9(1), 108–112.

Sawiah, S., Retnaningtyas, E., Siwi, R. P. Y., & Wulandari, A. (2024). Behavior Of Consuming Betel Nut On The Incidence Anemia In Pregnant Women At Posyandu Wasur Kampung, Working Area Of Rimba Jaya Health Center Merauke District-Merauke Regency. Journal of Health Science Community, 5(2), 127–133.

Senn, M., Baiwog, F., Winmai, J., Mueller, I., Rogerson, S., & Senn, N. (2009). Betel nut chewing during pregnancy, Madang province, Papua New Guinea. Drug and alcohol dependence, 105(1-2), 126–131.

Shenk, M. K., Morse, A., Mattison, S. M., Sear, R., Alam, N., Raqib, R.,… Shaver, J. (2021). Social support, nutrition and health among women in rural Bangladesh: complex tradeoffs in allocare, kin proximity and support network size. Philosophical Transactions of the Royal Society B, 376(1827), 20200027.

Singroha, K., & Kamath, V. V. (2016). Liver function tests as a measure of hepatotoxicity in areca nut chewers. Journal of Dental Research and Review, 3(2), 60–64.

Sohag, A. A. M., Hossain, M. T., Rahaman, M. A., Rahman, P., Hasan, M. S., Das, R. C.,… Uddin, M. J. (2022). Molecular pharmacology and therapeutic advances of the pentacyclic triterpene lupeol. Phytomedicine, 99, 154012.

Strickland, S., & Duffield, A. (1997). Anthropometric status and resting metabolic rate in users of the areca nut and smokers of tobacco in rural Sarawak. Annals of human biology, 24(5), 453–474.

Strickland, S., Veena, G., Houghton, P., Stanford, S., & Kurpad, A. (2003). Areca nut, energy metabolism and hunger in Asian men. Annals of human biology, 30(1), 26–52.

Sullivan, R. J., Allen, J. S., Otto, C., Tiobech, J., & Nero, K. (2000). Effects of chewing betel nut (Areca catechu) on the symptoms of people with schizophrenia in Palau, Micronesia. The British journal of psychiatry, 177(2), 174–178.

Sullivan, R. J., Andres D Ch MS, P. D. M., Sylvia, Otto, C., Miles, W., & Kydd, R. (2007). The effects of an indigenous muscarinic drug, Betel nut (Areca catechu), on the symptoms of schizophrenia: a longitudinal study in Palau, Micronesia. American Journal of Psychiatry, 164(4), 670–673.

Sun, H., Yu, W., Li, H., Hu, X., & Wang, X. (2024). Bioactive components of areca nut: an overview of their positive impacts targeting different organs. Nutrients, 16(5), 695.

Suzuki, K., Jayasena, C. N., & Bloom, S. R. (2012). Obesity and appetite control. Experimental diabetes research, 2012.

Sznajder, K., Wander, K., Mattison, S., Medina, E. A., Alam, N., Raqib, R.,… Shenk, M. K. (2021). Labor Migration Is Associated With Lower Rates of Underweight and Higher Rates of Obesity Among Left-behind Wives in Rural Bangladesh: a Cross-sectional Study.

Sznajder, K. K., Shenk, M. K., Alam, N., Raqib, R., Kumar, A., Haque, F.,… Wander, K. (2023). Betel quid use is associated with anemia among both men and women in Matlab, Bangladesh. PLOS Global Public Health, 3(6), e0001677.

The Syndey Morning Herald Reporter. (2008). Betelnut is good against diabetes: study. The Syndey Morning Herald.

Thomas, S. J., Harris, R., Ness, A. R., Taulo, J., Maclennan, R., Howes, N., & Bain, C. J. (2008). Betel quid not containing tobacco and oral leukoplakia: a report on a cross-sectional study in Papua New Guinea and a meta-analysis of current evidence. International Journal of Cancer, 123(8), 1871–1876.

Toprani, R., & Patel, D. (2013). Betel leaf: Revisiting the benefits of an ancient Indian herb. South Asian Journal of Cancer, 2(3), 140.

Tsai, W.-C., Wu, M.-T., Wang, G.-J., Lee, K.-T., Lee, C.-H., Lu, Y.-H.,… Lin, T.-H. (2012). Chewing areca nut increases the risk of coronary artery disease in Taiwanese men: a case-control study. BMC public health, 12, 1–7.

Tseng, C.-H. (2008). Betel nut chewing is associated with hypertension in Taiwanese type 2 diabetic patients. Hypertension Research, 31(3), 417–423.

Tseng, C.-H. (2010). Betel nut chewing and incidence of newly diagnosed type 2 diabetes mellitus in Taiwan. BMC research notes, 3(1), 1–8.

Tung, T.-H., Chiu, Y.-H., Chen, L.-S., Wu, H.-M., Boucher, B., & Chen, T.-H. (2004). A population-based study of the association between areca nut chewing and type 2 diabetes mellitus in men (Keelung Community-based Integrated Screening programme No. 2). Diabetologia, 47(10), 1776–1781.

VanderWeele, T. J., & Ding, P. (2017). Sensitivity analysis in observational research: introducing the E-value. Annals of internal medicine, 167(4), 268–274.

Wander, K., Brindle, E., & O’Connor, K. A. (2012). Sensitivity and specificity of C-reactive protein and α1-acid glycoprotein for episodes of acute infection among children in Kilimanjaro, Tanzania. American Journal of Human Biology, 24(4), 565–568.

Wang, L., & Hui, S. S.-c. (2015). Validity of four commercial bioelectrical impedance scales in measuring body fat among Chinese children and adolescents. BioMed Research International, 2015.

Warnakulasuriya, S., & Chen, T. (2022). Areca nut and oral cancer: evidence from studies conducted in humans. Journal of Dental Research, 101(10), 1139–1146.

Wei, Y.-T., Chou, Y.-T., Yang, Y.-C., Chou, C.-Y., Lu, F.-H., Chang, C.-J., & Wu, J.-S. (2017). Betel nut chewing associated with increased risk of arterial stiffness. Drug and alcohol dependence, 180, 1–6.

Weir, C. B., & Jan, A. (2019). BMI classification percentile and cut off points. StatPearls [Internet].

Wu, F., Parvez, F., Islam, T., Ahmed, A., Rakibuz-Zaman, M., Hasan, R.,… Ahsan, H. (2015). Betel quid use and mortality in Bangladesh: a cohort study. Bulletin of the World Health Organization, 93, 684–692.

Yamada, T., Hara, K., & Kadowaki, T. (2013). Chewing betel quid and the risk of metabolic disease, cardiovascular disease, and all-cause mortality: a meta-analysis. PloS one, 8(8), e70679.

Yang, M.-S., Lee, C.-H., Chang, S.-J., Chung, T.-C., Tsai, E.-M., Ko, A. M.-J., & Ko, Y.-C. (2008). The effect of maternal betel quid exposure during pregnancy on adverse birth outcomes among aborigines in Taiwan. Drug and alcohol dependence, 95(1-2), 134–139.

Yang, N.-Y. J., Kaphle, K., Wang, P.-H., Jong, D.-S., Wu, L.-S., & Lin, J.-H. (2004). Effects of aqueous extracts of" betel quid" and its constituents on testosterone production by dispersed mouse interstitial cells. The American Journal of Chinese Medicine, 32(05), 705–715.

Zaw, K. K., Ohnmar, M., Hlaing, M. M., Oo, Y. T., Win, S. S., Htike, M. M.,… Thein, Z. M. (2016). Betel Quid and Oral Potentially Malignant Disorders in a Periurban Township in Myanmar. PloS one, 11(9), e0162081. doi:10.1371/journal.pone.0162081

